# Resting State Functional Connectivity Alterations in Individuals with Autism Spectrum Disorders: A Systematic Review

**DOI:** 10.1101/2021.07.27.21261165

**Authors:** Varun Kumar, Rahul Garg

## Abstract

Many studies investigate the alterations in resting state functional connectivity in autism. Most of these studies focus on different regions of the brain to find the connectivity differences between autism spectrum disorder and typically developing populations. The present review quantitatively extracts this data from all the seed based studies on autism spectrum disorder and uses it to build, for the first time, an Autism Altered Functional Connectome (AAFC) which summarizes the alterations in functional connectivity consistently reported in the literature. The data extracted from all the studies matching the inclusion criteria are presented at one place in human as well as a machine-readable format for further interpretation and analysis. Systematically reviewing 41 publications on 2818 subjects comprising 1459 typically developing and 1359 subjects with autism spectrum disorder, a total of 932 altered functional connectivity links were employed to construct an AAFC. The AAL atlas mapping of these links resulted in 71 replicated links of which 49 were consistent, and 574 unreplicated links that were reported just once. **Out of 49, 38 were replicated across different non overlapping datasets**. Majority of the studies analyzed the functional connectivity of the Default Mode Network (DMN) and its regions. Two important DMN regions, namely precuneus and posterior cingulate cortex were reported to exhibit different connectivity profiles with former majorly underconnected and later majorly overconnected consistently reported across multiple studies. After mapping the AAFLs to an atlas of brain networks, poor integration within DMN regions, and poor segregation of DMN regions with extra-DMN regions was observed.

## 1. Introduction

Diagnostic and Statistical Manual of Mental Disorders 5*^th^* edition (DSM-V) defines Autism spectrum disorder (ASD) as a neurodevelopmental disorder characterized by impaired social interaction and the display of restricted and repetitive patterns of behaviors and interests (American Psychiatric Association, 2013). ASD is not a single disorder but comprises a spectrum of disorders. A person with ASD shows a range of developmental delays ranging from limited and repetitive behaviors to developing strong interests in some field but being socially impaired. Centers for Disease Control and Prevention (CDC) in their study (Baio et al., 2018) involving eight-year-old children from 11 sites in the United States found that in the year 2014 the prevalence rate of ASD was 1 in 59 children. Presently, there is no known cure for ASD (Russell et al., 2018). The patient is given behavioral therapies to improve behavior and speech. Sometimes the medications are given to cure the comorbidities. The treatments are most effective when given at an early age (Elder et al., 2017). The high prevalence rate together with the fact of ASD being noncurable till now brings upon the urgent need to find the cause as well as the most effective treatment or management strategies to handle this condition at an early age. It is hoped that finding out brain mechanisms disrupted in ASD will give pointers to its etiology as well as the treatment strategies.

The functional neuroimaging based on fMRI provides a non-invasive means to understand brain function. Functional MRI measures blood oxygenation level dependent (BOLD) contrast of the brain. The BOLD signal acts as a surrogate neuronal signal by measuring the changes in the blood flow possibly triggered by changes in the neuronal firing patterns. This technology could pave the way to identify the brain processes uniquely disrupted in ASD.

Historically, the methods to find brain regions activated in response to a specific task or condition (or differently activated under different conditions or tasks), required task-based experimental protocol design (Frackowiak et al., 2004). The fMRI data thus acquired was analyzed using the General Linear Model (GLM) analysis method. The identified the brain regions that activated differently among individuals with ASD and typically developing (TD) individuals under the specific experimental condition studied. Such studies start with an apriori hypothesis about disruptions in brain function (such as processing or recognition of human faces) and design the experimental protocols to elicit responses in specific areas of the brain (for example, showing images of human faces versus crosshair). Various task-based fMRI activation studies on ASD have investigated the brain regions involved in emotional processes (Maddock, 1999) self-referential thought (Northoff and Bermpohl, 2004) and Theory of Mind (Ochsner et al., 2005), which are impaired in ASD (Frith, 2003).

The task of selective attention and cognitive flexibility (Kennedy et al., 2006) and activation in the fusiform gyrus during a face stimulus (Koshino et al., 2007), (Pierce and Redcay, 2008) have also been investigated. See (Minshew and Keller, 2010), (Philip et al., 2012) for a comprehensive review on task-based fMRI studies on ASD.

In recent years, especially after the discovery of default mode network (DMN) in brain (Raichle et al., 2001), (Hutchison et al., 2013) there has been a growing interest in understanding the interactions among various brain regions.

These interactions have been characterized by a measure of correlations of BOLD signals between different brain regions, both in the task-based and resting state paradigm (which requires the participant to just lie down in the scanner and not to think about anything in particular). It was found that there are regions in the brain (now known as default mode network or DMN) that get deactivated on the presentation of attention-demanding task (Shulman et al., 1997). It was also discovered that the sub-regions of DMN exhibit a very high functional correlation of BOLD signals, among themselves, even in the state of rest (during resting state paradigm). Since then several brain networks have been discovered in the resting state paradigm based on the correlations of the BOLD signal among different brain regions (Schaefer et al., 2017), (Damoiseaux et al., 2006).

The study of interactions among different brain regions is commonly referred to as connectivity analysis. The studies analyzing connectivity can be classified into doing (a) structural connectivity analysis, which aims to find out anatomically laid out neuronal fibers or fiber tracts among different brain regions (typically using imaging modalities such as Diffusion Tensor Imaging (DTI) (Le Bihan et al., 2001); (b) functional connectivity analysis, which characterizes the interactions among various brain regions by correlations in BOLD time series (van den Heuvel and Pol, 2010), (Smith, 2012); and (c) effective connectivity analysis, which aims to find ‘causal’ interactions among different brain regions while removing the confounding effect of other regions (Friston, 2011). While the effective connectivity methods are still evolving, functional connectivity analysis is becoming very successful in finding out disruptive brain networks for various mental disorders such as Alzheimer’s disease, depression, and schizophrenia (Greicius, 2008), (Greicius et al., 2004), (Garrity et al., 2007).

Several large scale projects such as human connectome project (Essen et al., 2013), 1000 functional connectomes project (Biswal et al., 2010) and NKI-Rockland Sample (Nooner et al., 2012) have been launched to map all the functional networks in healthy human subjects. Also, large scale repositories of freely accessible data such as ABIDE-I (Di Martino et al., 2013) and ABIDE-II (Di Martino et al., 2017) have been created to study disruptions in functional connectivity in ASD.

There have been many studies that reviewed the task-based fMRI studies in ASD (Philip et al., 2012, Vissers et al., 2012, Minshew and Keller, 2010, Wass, 2011, NJ and DL, 2007, Anagnostou and Taylor, 2011, Maximo et al., 2014, Pina-Camacho et al., 2012, Harms et al., 2010, Sanders et al., 2008, Williams and Minshew, 2007).

Similarly, multiple reviews, majority of them being narrative (Jack, 2018, Hull et al., 2017, Li et al., 2017, Crippa et al., 2016, D’Mello and Stoodley, 2015, Uddin et al., 2013), investigated the studies that reported disruptions disruptions in the functional connectivity in the resting state paradigm.

Rane et al. (Rane et al., 2015) systematically reviewed the studies based on resting state fMRI as well as DTI and concluded that majorly, the long-range functional connectivity decreases in autistic individuals as compared to typically developing. Muller et al. (Müller et al., 2011) in their systematic review tried to find methodological differences between studies that reported only underconnectivity and the studies that reported a mix of under and over connectivity (or just overconnectivity).

The reviews on resting state paradigm have summarized the data about functional connectivity alterations between brain regions in the form of tables using the region names as reported in the studies that were investigated. Different studies use different atlases to report the region names corresponding to their findings. Use of different atlases may give different or inconsistent results, especially when the number of voxels in a differently connected cluster is small and the cluster lies at the boundary of two regions. For example, the MNI coordinates (-39, -17, 42) lie in left precentral gyrus according to Talairach-Tournoux atlas, whereas they lie in the left postcentral gyrus according to the CA_ML_18_MNIA atlas (in AFNI). Thus the results reported may become slightly inconsistent. This inconsistency will be discussed in detail in section 2.7.

The present review goes a step further and attempts to unambiguously describe changes in resting-state functional connectivity (rc-FC) in individuals with ASD by systematically reviewing the studies on 2818 subjects comprising 1459 typically developing and 1359 subjects with ASD. The review constructs an Autism Altered Functional Connectome (AAFC), using all the past studies on rs-FC in ASD individuals, meeting the inclusion/exclusion criteria. The AAFC consists of a collection of altered functional connectivity links between pairs of brain regions. The brain regions are represented by their corresponding coordinates in standard Montreal Neurological Institute (MNI) space. The altered functional connectivity links may be underconnected in individuals with ASD, implying that the correlation between the two regions is significantly less (either more negative or less positive) in the ASD population as compared to typically developing population. Similarly, an overconnected functional connectivity link means that the corresponding correlation in ASD population is significantly higher in ASD as compared to TD population.

Thus, the review extracts the data on connectivity alterations from all the rs-FC studies on ASD employing seed ROI based analysis methodology and converts it into a collection of links summarizing the results of all the research in rs-FC of ASD. It then maps this data back to brain regions using different atlases to find replicated, consistent and inconsistent altered functional connectivity links. These replicated links are labeled as consistent if all the studies replicating the link find it under or overconnected. They are labeled as inconsistent if some studies find it underconnected while others find it overconnected. These links are mapped onto three atlases to get the above replication statistics at different levels of granularity. They are also mapped to brain lobes and different functional networks to find overall patterns of altered functional connectivity in ASD.

Forty-one publications on rs-FC differences in ASD were found meeting our inclusion/exclusion criteria. From these publications, a total of 932 altered functional connectivity links were extracted and analyzed to construct the AAFC. Mapping these 932 links onto the AAL atlas (Tzourio-Mazoyer et al., 2002), translated it to unique 645 links of AAL regions out of which 71 links were reported each by at least two studies. These 71 unique replicated links corresponded to 49 consistent links and 22 inconsistent links. Out of total 645 unique links mapped to AAL atlas, 574 links appeared in single studies only, which suggests that the present focus of most of the autism research in seed ROI based rs-FC analysis is on fresh discovery rather than refinements or replications. With the availability of large datasets such as ABIDE-I and II, the trend is expected to shift more to refinements and replications.

After mapping the links to AAL atlas, posterior cingulate cortex (PCC) was observed to be overconnected with multiple brain regions and precuneus to be underconnected with multiple brain regions. During the lobe level analysis, the cerebellum was observed to be majorly underconnected with the frontal lobe, and occipital lobe to be overconnected to the limbic lobe in ASD individuals as compared to TD. After mapping the links to functional networks, the regions of DMN were observed to be majorly underconnected within themselves and majorly overconnected to the extra-DMN regions. A major cause of inconsistencies was observed to be the gender as well as the large anatomical distance between the endpoints of a pair of links.

The consistent connectome hence discovered is available in the form of a spreadsheet in supplementary information along with other details such as preprocessing parameters, nuisance covariates removed, subjects’ aggregate demographics, of the studies reviewed. This along with other tables and figures in this review may serve as a valuable resource for researchers in the field of ASD. They may now, not need to go through individual seed ROI based rs-FC study in ASD and can directly examine and analyze the data in the spreadsheet, and may accelerate the discovery of neural mechanisms involved in ASD.

This study puts all information on seed ROI based resting-state functional connectivity changes in ASD at one place in a structured format and reports the consistencies and inconsistencies in the published literature. Moving form study oriented description to region-pairs oriented description of alterations, this review gives a machine-readable, quantitative format to query if a pair of regions demonstrates altered connectivity in ASD.

The paper is organized as follows. Section 2 discusses the methods employed to search and shortlist relevant publications. It then describes the details of the methodology employed to extract the data from the publications selected for review to construct the AAFC.

Section 3 presents the results obtained after the literature search, data extraction and mapping the extracted data onto various brain atlases. Section 4 interprets the consistencies and inconsistencies observed in the review. Section 5 concludes the study by focusing on the major contributions of the study and pointing to the future work.

## 2. Methods

### 2.1. Background: Resting State Functional Connectivity Analysis

The functional connectivity analysis can be done for task-based experimental paradigms as well as resting state paradigms. Task-based studies require the participant to engage in a task. The task may be active such as speaking, moving a part of their body, etc. or passive such as listening or passive viewing. Resting state studies require the participant to lie down in the scanner and not to think about anything in particular. Some studies ask them to close their eyes, while some to keep the eyes open and focus on a crosshair. As the experimental paradigm followed in resting state experiment across studies is quite similar, the scans from various sites can be pooled together to construct a bigger data set. Also, the replication is easier to achieve as the experimental paradigm is simple and the same for many studies. It is difficult to scan atypically developing children due to multiple reasons such as excessive head motion, refusal to comply with the task and inattention (Yerys et al., 2009). As resting state paradigm needs very less in-scanner compliance with the participant, it is heavily used with young as well as diseased population such as ASD. Therefore this review focuses on rs-FC paradigm based studies in ASD.

The functional connectivity analysis can be carried out at levels of (i) Seed ROI based; (ii) component based; and (iii) global graph theory based. The seed ROI based functional connectivity (FC) analysis is the most common analysis method (Hull et al., 2017). It starts with defining spatially contiguous regions of interest (ROIs). These ROIs may either be defined by experts or may be selected from a standard atlas. After defining the ROIs, the changes in functional connectivity can be studied at the level of ROI-to-ROI (also called ROI-based or RB), in which functional connectivity is calculated between the representative time courses of two ROIs, or ROI-to-voxels or seed-to-voxels (also called seed-based or SB), in which the functional connectivity is calculated between the representative time courses of one ROI (also called seed) and all the other brain voxels. In component-based analysis, instead of spatially contiguous ROIs, several network components are studied. These components, typically calculated using data-driven methods such as Independent Component Analysis (ICA), may not be contiguous and may also be overlapping with other components. The functional connectivity analysis can be done among components (by calculating mean time courses of these components) or from component to voxels. The graph-theoretic analysis considers several global graph theoretic measures of connectivity graphs.

### 2.2. Inclusion Criteria

The inclusion/exclusion criteria is as follows. (i) Experimental paradigm: Any task-based study was excluded from the review. Only studies analyzing the resting state paradigm was considered irrespective of the eye status of the study participants during scan. (ii) Analysis method: This review focuses on studies that used seed ROI based functional connectivity analysis on resting state data. So, the studies employing noncontiguous ROIs (using ICA or other methods) or graph-theoretic analysis, were excluded. (iii) Study group: This review includes studies that have investigated group differences of ASD and TD. Studies on mice or just on a single group of healthy/ASD individuals were excluded. (iv) Review articles were excluded. (v) Studies which had not performed the correction for multiple comparisons were excluded. (vi) Coordinates: Studies for which the seed, target or ROI coordinates could not be estimated were excluded. (vii) Articles not published in English were excluded. (viii) The articles that just investigated the changes in the hemispheric connectivity for a seed (i.e. how the connectivity between bilateral seeds change among themselves) were not included. (ix) Studies that regressed out task effects from the fMRI data were not included.

### 2.3. Search Strategy and Study Section

The systematic review was conducted on the PubMed articles up to 9*^th^*April 2018 according to the standard - Preferred Reporting Items for Systematic Reviews and Meta-Analysis Guidelines (PRISMA) (Liberati et al., 2009). The search query included terms such as fMRI, resting state, autism and connectivity. Various negated (NOT) terms were also included in conjunction with the query such as MEG, task-based and independent component analysis to exclude studies of no interest. Refer to the Appendix A.1 for the detailed query and its explanation.

The search query resulted in unique 347 publications. The results were further screened for inclusion as described as follows.

The publications resulted from the PubMed went through two steps of screening. In the first step, the publications were selected by reading just the title and the abstract. This resulted in an exclusion of 263 publications. The remaining 84 publications went through the second step of screening. This was done by reading the entire paper and including the ones that satisfied the inclusion/exclusion criteria mentioned in section 2.2. Thirty-seven publications failed the inclusion criteria, and six publications were excluded during data extraction. Finally, 41 publications were included in the review. The Figure 1 shows a pictorial representation of the study selection process.

**Figure 1:**
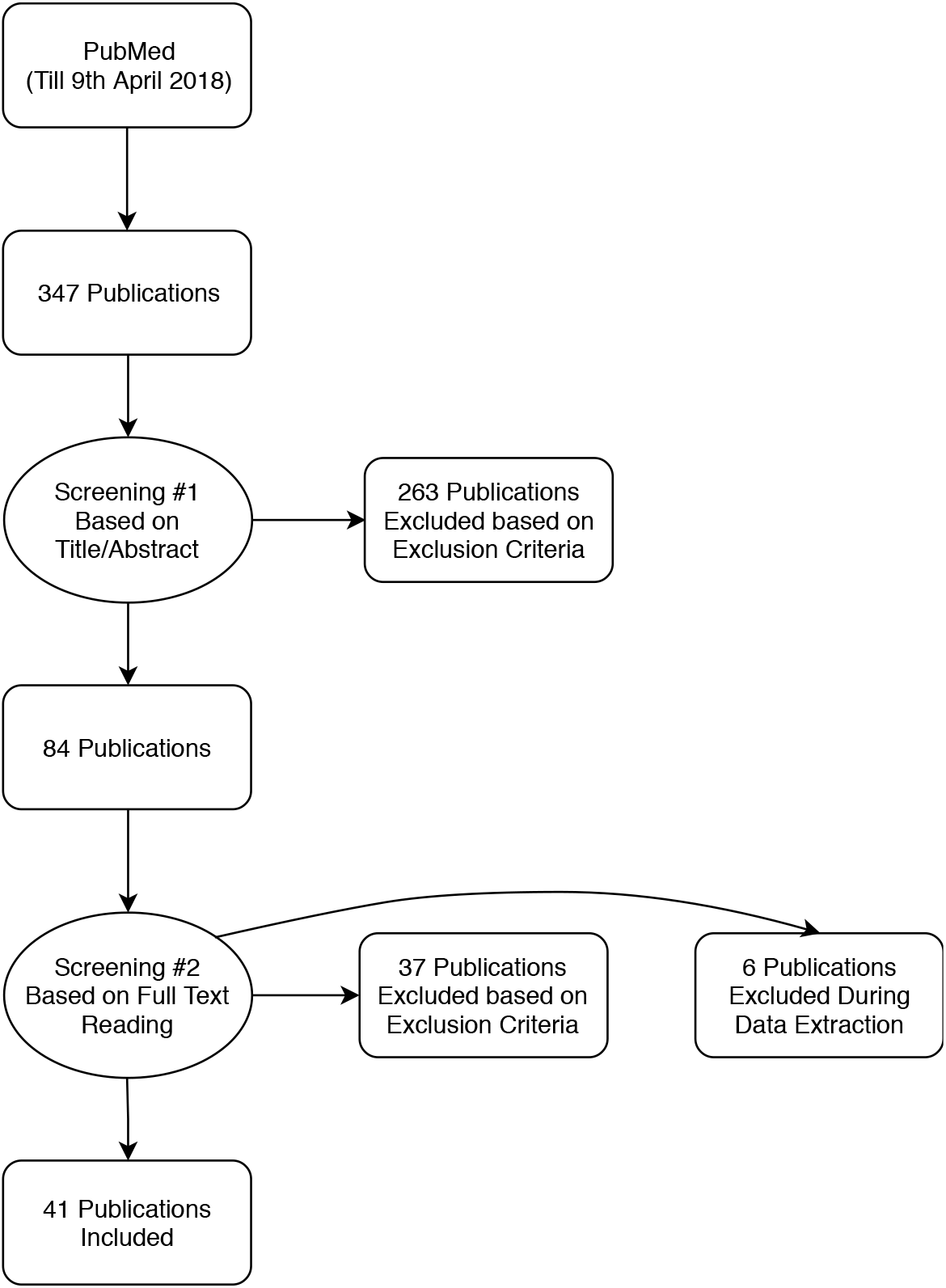
Flowchart depicting the study selection process for the present systematic review.

### 2.4. Definitions

#### Publication

For this review, a publication is defined as a peer-reviewed article meeting the inclusion/exclusion criteria defined in section 2.2. A total of 41 publications have been reviewed in this article.

#### Study

If a publication involved independent investigation two different non-overlapping samples of subjects, it is said to be composed of two studies. Otherwise, it is stated to be composed of one study. For example, separate male and female cohorts in (Alaerts et al., 2016), two different sites analyzed independently by (Guo et al., 2016), and independent analysis of two cohorts - children and adults by (Burrows et al., 2016), each consists of two studies. Therefore the 41 publications translated to 44 studies. Each study is being given an ID (the ‘Study’ column in the Table 1) that is then referred to in the main text.

**Table 1.**
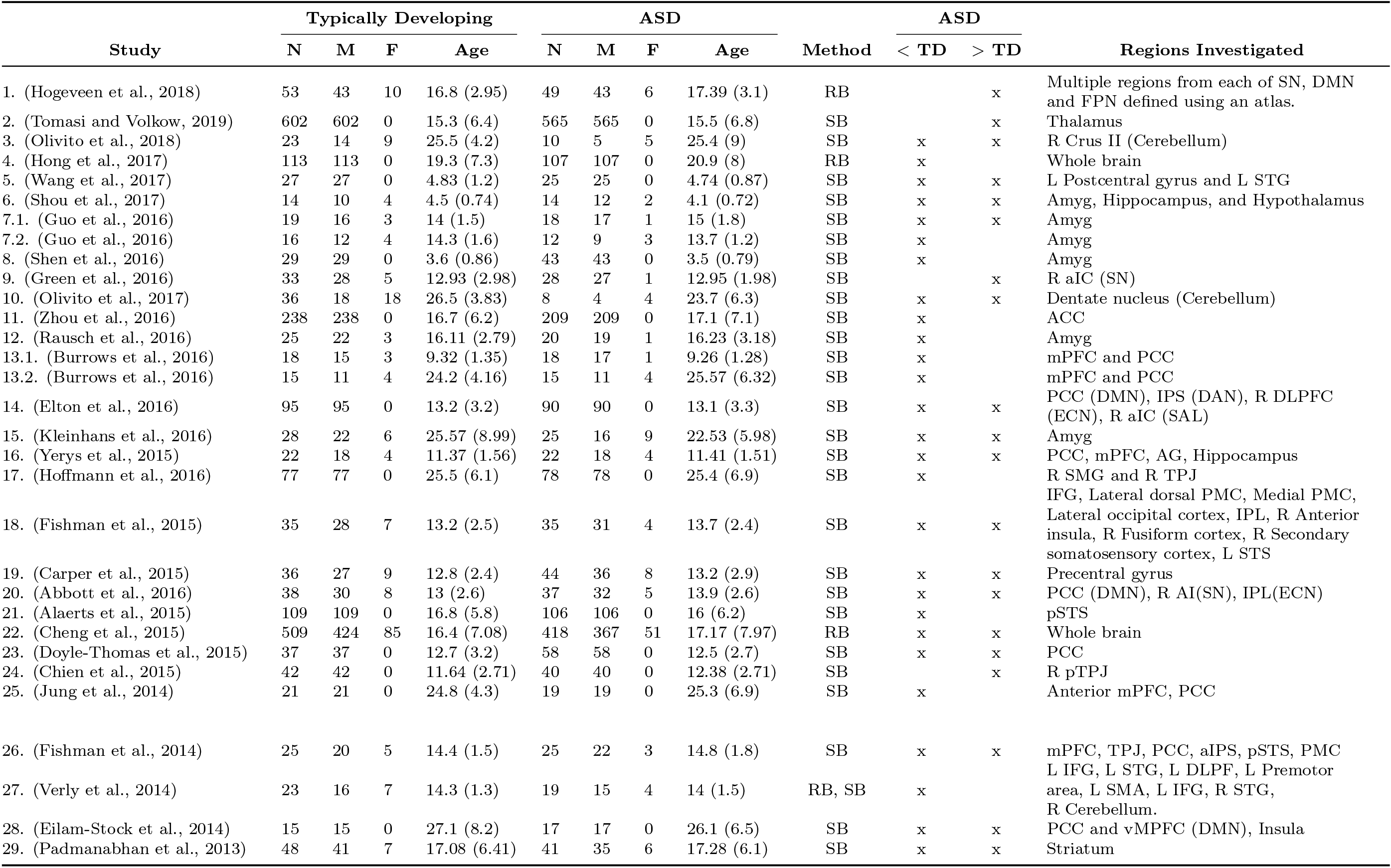
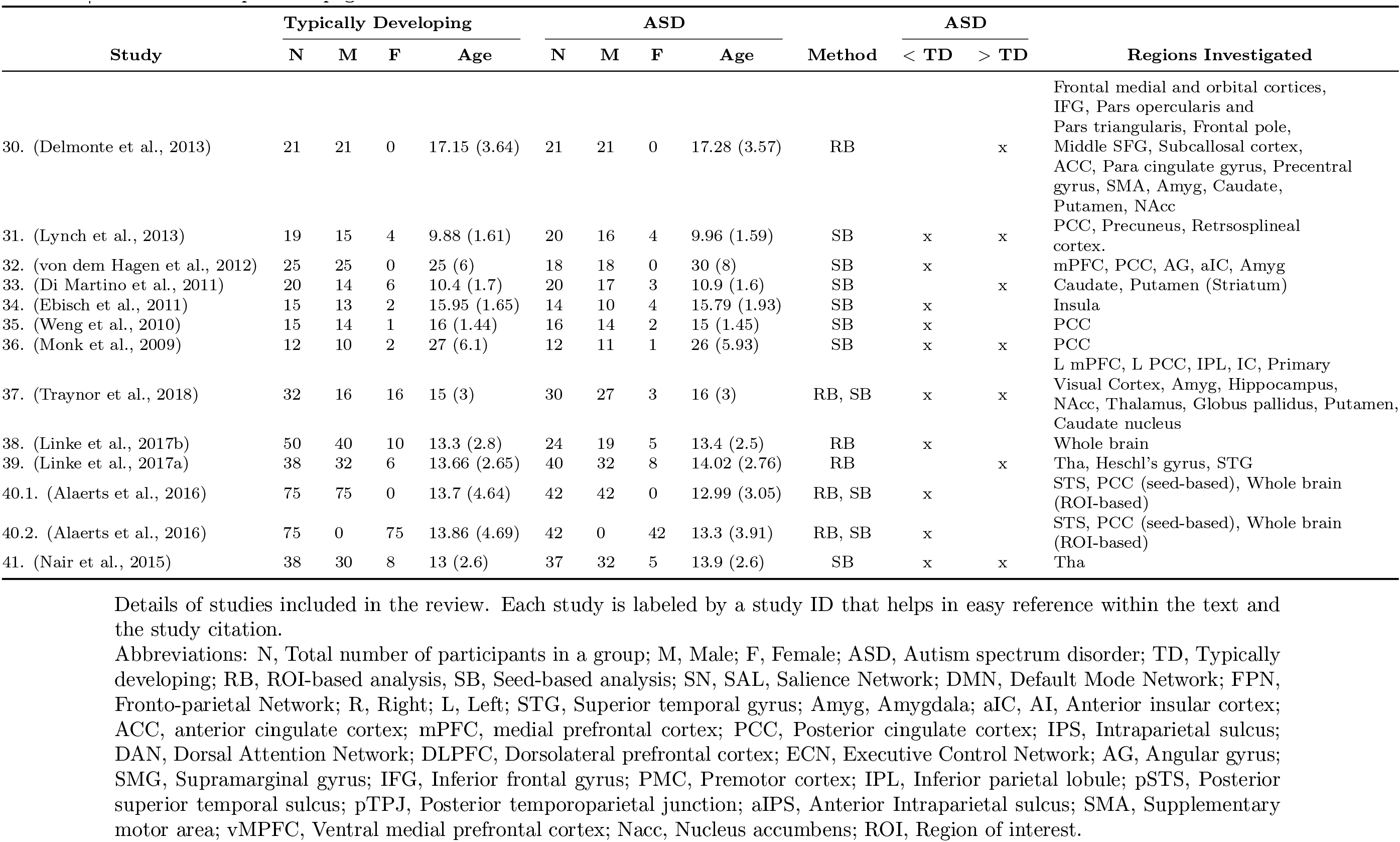
Details of studies included in the review

#### Autism Altered Functional Connectivity Link (AAFL)

AAFL represents a pair of brain regions along with the altered connectivity description between them. The brain regions are represented by their corresponding coordinates in standard MNI space. An AAFL may be underconnected in individuals with ASD, implying that the correlation between the two regions is significantly less (either more negative or less positive) in the ASD population as compared to typically developing population. Similarly, an overconnected AAFL means that the corresponding correlation in ASD population is significantly higher in ASD as compared to TD population. In this review, the term ‘link’ will be used synonymously with AAFL for the ease of reading.

#### Replicated Link

An AAFL reported in a study is said to be replicated with respect to an atlas, if there exists at least one more study reporting another AAFL such that MNI coordinates of both the links correspond to the same regions in the atlas. For example, one study may report a link (-29,-6,1) underconnected to (24,-6,-12) whereas another study may report a link (-25,-3,3) overconnected to (21,-3,-11). Mapping the coordinates to Harvard Oxford subcortical atlas, the coordinates (-29,-6,1) and (-25,-3,3) both belong to the putamen, and (24,-6,-12) and (-25,-3,3) both belong to the amygdala. Thus the (unique) link between putamen and amygdala is replicated with respect to Harvard Oxford subcortical atlas. Similarly, the above mentioned unique link would also be called replicated if, without loss of generality, the endpoints of the second link were swapped.

#### Consistent Link

A replicated link is termed as consistent if it has the same connectivity (under-connectivity or over-connectivity) across all the studies it is reported in.

#### Inconsistent Link

A replicated link is said to be inconsistent some of the studies report overconnectivity while others report underconnectvity for it.

#### Perfect Replicated Link

A perfectly replicated link is the link that is reported to have same connectivity by at least two studies that analyzed different non-overlapping datasets. A link cannot be called perfectly replicated if, for example, all the studies reporting that link have used the ABIDE dataset. It is to be noted that not all replicated links are perfectly replicated.

### 2.5. Altered Functional Connectome

An Altered Functional Connectome (AFC) consists of a collection of consistently replicated links between pairs of brain regions. After the atlas mapping, statistical analysis could be carried out to find the altered functional connectivity links. Such an analysis will be similar to the multilevel kernel density analysis (MKDA) (Wager et al., 2009), except that unlike the current applications, for every seed region, a separate MKDA analysis needs to be carried out to construct the altered functional connectome. Such an analysis will result in a meta-study systematically consolidating all past the research on altered resting-state functional connectivity in ASD. For suitable statistical power, one will require a reasonable number (say 10-15) of studies reporting connectivity difference between the same seed-target pairs. Currently, the state of ASD research is more focused on discovery and less on replication of earlier results (see section 3.3). The number of studies reporting connectivity difference between the same pairs of regions is unacceptably small for a meta-study, even after mapping the MNI coordinates to a coarse atlas such as the AAL atlas (Tzourio-Mazoyer et al., 2002). It was possible to carry out a meta-analysis by mapping the seed coordinates to a large network such as the default mode network (DMN) to the salience network (SN) as done by (Kaiser et al., 2015), (Sha et al., 2019). Statistical analysis methods such as ALE (Eickhoff et al., 2009), KDA (Etkin and Wager, 2007) or MKDA (which overcomes the limitations of other approaches) (Kober and Wager, 2010) may be applied to get the final results. However, such an approach must be applied with extreme caution as it may lead to incorrect results. To see this, consider the hypothetical scenario of Figure 2(A) with two subregions (R11 and R12) of the network N1 having different connectivity to a voxel V1. Imagine ten links between R11 and voxel V1 to be reported in the literature as underconnected, and ten links as overconnected. In this case, the MKDA method with aggregation applied at the level of network N1 will result in an inconclusive finding between the N1 and V1, since the statistical analysis will find ten overconnected and ten underconnected links between N1 and V1. The AFC approach suggested earlier in this section will correctly conclude region R11 to be underconnected and region R12 to be overconnected to V1 after the statistical analysis. In another study (Sha et al., 2019) the reported seed and peak coordinates were mapped to an atlas of 1024 parcellations and statistical analysis was done independently for underconnected and overconnected links. Such an approach of analyzing the overconnected and underconnected links separately may also lead to incorrect results. To see this, consider the hypothetical situation depicted in Figure 2(B), where regions R1 and R2 are reported to be overconnected and underconnected by ten studies each. The analysis method of (Sha et al., 2019) would conclude that regions R1 and R2 are both overconnected as well as underconnected, whereas the correct result (as found by proposed method) should have been no conclusive evidence alterations in functional connectivity between R1 and R2.

**Figure 2:**
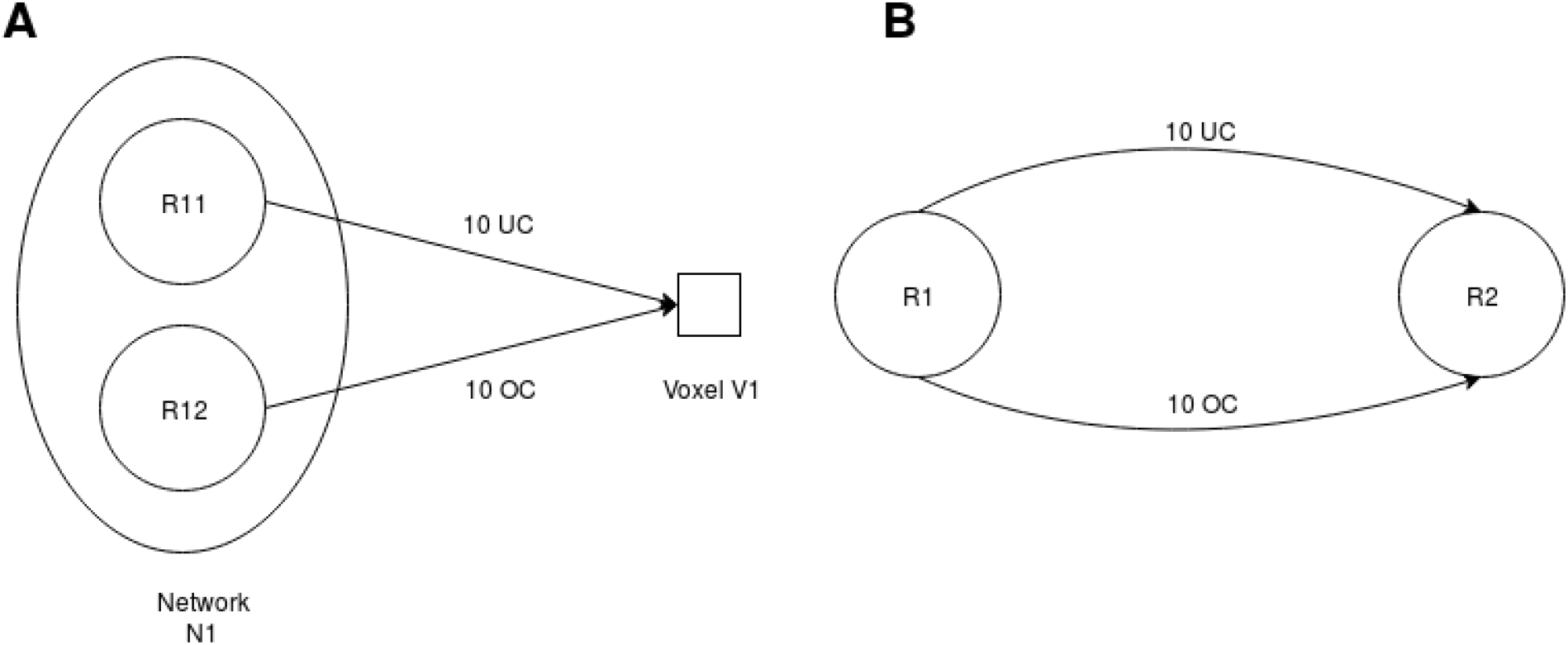
Scenarios showing the limitations of traditional statistical analysis approaches. Abbreviations: R1, Region1; R2, Region 2; UC, Underconnectivity; OC, Overconnectivity.

Due to these limitations, the present review, instead of carrying out a meta-study, limits itself to a quantitative systematic review and computes the Autism Altered Functional Connectome (AAFC) by using consistently replicated AAFLs. The inconsistent links, though not a part of AAFC are also reported for completeness and further analysis.

### 2.6. Data Extraction

To build an AAFC, the first step was to extract the MNI coordinates as well as the connectivity information mentioned in the results of the studies in a consistent manner.

For ROI-to-voxels based studies, the coordinates of the seed ROIs and the cluster’s peak value voxels were extracted. For ROI-to-ROI based studies coordinates of ROIs were extracted. If the coordinates were not reported, the same atlas used by the study was employed to get the MNI coordinates of the center of mass of ROIs. If the center of mass laid outside the region, then the voxel in the corresponding ROI closest to the center of mass was selected. In case the coordinates could not be estimated, the authors were contacted over email. We were successful in receiving coordinates of two studies (Hogeveen et al., 2018), (Hong et al., 2017). A web application (Papademetris, 2018) based on the paper by Lacadie et al. (Lacadie et al., 2008) was used to convert the coordinates to MNI in case they were reported in Talairach space. The regions for which the coordinates could not be estimated were discarded. In addition to the coordinates, various preprocessing details, as well as demographic details, were also extracted as shown in the Tables 1 and 2.

**Table 2.** Preprocessing Details

| Study | Highpass<br>(Hz) | Lowpass<br>(Hz) | Smooth<br>(fwhm)<br>mm | GSR | CSF | WM | Motion<br>Params | Scrub | GCOR | CR |
| --- | --- | --- | --- | --- | --- | --- | --- | --- | --- | --- |
| 1. (Hogeveen et al., 2018) | 0.01 | 0.1 | 6 |  | x | x |  | x | x |  |
| 2. (Tomasi and Volkow, 2019) | 0.01 | 0.1 | 4 | x | x | x | x | x |  |  |
| 3. (Olivito et al., 2018) | 0.01 | 0.08 | 8 | x |  |  | x |  |  |  |
| 4. (Hong et al., 2017) | 0.01 | 0.1 | 5 |  | x | x | x |  |  |  |
| 5. (Wang et al., 2017) | 0.01 | 0.1 | 6 |  | x | x | x | x |  |  |
| 6. (Shou et al., 2017) | 0.01 | 0.08 | 8 | x | x | x | x |  |  |  |
| 7. (Guo et al., 2016) | 0.01 | 0.08 | 8 | x | x | x | x | x |  |  |
| 8. (Shen et al., 2016) | 0.008 | 0.08 | 6 | x | x | x | x | x |  |  |
| 9. (Green et al., 2016) | 0.01 | 0.1 | 5 | x | x | x |  | x |  |  |
| 10. (Olivito et al., 2017) | 0.01 | 0.08 | 8 | x |  |  | x |  |  |  |
| 11. (Zhou et al., 2016) | 0.01 | 0.08 | 8 |  | x | x | x |  |  |  |
| 12. (Rausch et al., 2016) | 0.01 | - | 6 |  | x | x | x |  |  | x |
| 13. (Burrows et al., 2016) | 0.01 | 0.08 | 5 |  | x | x | x | x |  |  |
| 14. (Elton et al., 2016) | 0.01 | 0.1 | - |  | x | x | x |  | x |  |
| 15. (Kleinhans et al., 2016) | 0.01 | - | 5 |  | x |  | x |  |  | x |
| 16. (Yerys et al., 2015) | 0.01 | 0.08 | 7 | x | x | x | x |  |  |  |
| 17. (Hoffmann et al., 2016) | 0.01 | 0.08 | 8 |  | x | x | x | x |  |  |
| 18. (Fishman et al., 2015) | 0.008 | 0.08 | 6 |  | x | x | x | x |  |  |
| 19. (Carper et al., 2015) | 0.008 | 0.08 | 6 |  | x | x | x | x |  |  |
| 20. (Abbott et al., 2016) | 0.008 | 0.08 | 6 |  | x | x | x | x |  |  |
| 21. (Alaerts et al., 2015) | 0.009 | 0.08 | 6 |  | x | x | x |  |  |  |
| 22. (Cheng et al., 2015) | 0.01 | 0.08 | 8 | x | x | x | x | x |  |  |
| 23. (Doyle-Thomas et al., 2015) | 0.01 | 0.2 | 7 | x | x | x | x | x |  |  |
| 24. (Chien et al., 2015) | 0.01 | 0.08 | 8 |  | x | x | x |  |  |  |
| 25. (Jung et al., 2014) | 0.01 | 0.08 | 6 | x | x | x | x |  |  |  |
| 26. (Fishman et al., 2014) | 0.008 | 0.08 | 6 |  | x | x | x | x |  |  |
| 27. (Verly et al., 2014) | 0.009 | 0.08 | 8 |  | x | x | x |  |  |  |
| 28. (Eilam-Stock et al., 2014) | 0.01 | 0.08 | 8 | x | x | x | x |  |  |  |
| 29. (Padmanabhan et al., 2013) | 0.009 | 0.08 | 5 |  | x | x | x | x |  |  |
| 30. (Delmonte et al., 2013) | 0.008 | 0.2 | 5 |  | x | x | x |  |  |  |
| 31. (Lynch et al., 2013) | 0.008 | 0.1 | 6 | x | x | x | x |  |  |  |
| 32. (von dem Hagen et al., 2012) | 0.0078 | 0.08 | 8 | x |  |  | x |  |  |  |
| 33. (Di Martino et al., 2011) | 0.009 | 0.1 | 6 | x | x | x | x |  |  |  |
| 34. (Ebisch et al., 2011) | 0.009 | 0.08 | 6 | x | x | x | x |  |  |  |
| 35. (Weng et al., 2010) | - | 0.08 | 8 |  |  |  | x |  |  | x |
| 36. (Monk et al., 2009) | - | 0.08 | - |  |  |  |  |  |  | x |
| 37. (Traynor et al., 2018) | 0.008 | 0.09 | 6 |  | x | x | x | x |  |  |
| 38. (Linke et al., 2017b) | 0.008 | 0.08 | 6 |  | x | x | x | x |  |  |
| 39. (Linke et al., 2017a) | 0.008 | 0.08 | 6 |  | x | x | x | x |  |  |
| 40. (Alaerts et al., 2016) | 0.009 | 0.08 | 5 |  | x | x | x | x |  |  |
| 41. (Nair et al., 2015) | 0.008 | 0.08 | 6 | x | x | x | x | x |  | x |

Preprocessing details and the confounds regressed out. Each study is labeled by a study ID that helps in easy reference within the text and the study citation. Note that study ID 41 Nair et al. (2015) partialled out the whole cerebral cortex except the ROI under study. This is very similar to GSR. Therefore in the GSR column, this study is mentioned to have applied it. Abbreviations: High, Highpass filter range; Low, Lowpass filter range; Hz, Hertz; Smooth, Smoothing; fwhm, Full width half maximum; mm, millimeter; GSR, Global signal regression; CSF, Cerebrospinal Fluid; WM, White Matter; Scrub, Scrubbing; GCOR, Global correlation; CR, Cardiac and respiratory signals; ROI, Region of interest.

### 2.7. Atlas Mapping and Links Analysis

The replicated links cannot be found by the MNI coordinates of the extracted links alone as they have very fine granularity. It is very unlikely that two studies would report the exact same MNI coordinates for both the endpoints of an altered functional connectivity link. For determining the seed ROIs, different studies use different atlases. This results in significant variability in the reported MNI coordinates, even though the corresponding ROIs may be highly overlapping. In ROI-to-voxels analysis, the target ROIs are typically reported by a clustering tool after suitably thresholding the statistical maps. Thus, these ROIs are subjected to variabilities due to data, analysis methods and tools used.

To find replicated links, all the reported MNI coordinates of AAFL are converted back to region names using a brain atlas. Now, a link is said to be replicated if there is another study reporting another link with endpoints in the same two brain regions. See the section 2.4 for a more formal definition. Note that the replicated links are dependent on the choice of the brain atlas used. Using a fine-grained atlas is likely to result in a smaller number of replicated links as compared to a coarser brain atlas.

Note that the names of brain regions as reported in these studies cannot be used to create the consistent autism altered connectome. Different studies use different atlases to select the seed regions and to report the significantly connected voxels/clusters. But, different atlases are not perfectly consistent with each other. For a given MNI coordinate, different atlases can report different regions. For example, in a study by Elton et al. (Elton et al., 2016), the authors reported the coordinates (-15, -75, 48) as precuneus according to Talairach-Tournoux atlas whereas other atlases - Harvard Oxford atlas (in FSL) reports this coordinate as lateral occipital cortex, superior division and CA_ML_18_MNIA atlas (in AFNI) reports it as left superior parietal lobule. Therefore using just the region names to come up with a general connectome describing consistencies and inconsistency would lead to ambiguity. That is why, in the present review, first the MNI coordinates are extracted, and then they are mapped to an atlas for finding replicated links.

Three standard brain atlases of varied granularity - (1) composite Brainnetome Brain atlas (regions) (Fan et al., 2016) & Probabilistic Cerebellar atlass (Diedrichsen et al., 2009), which was of highest granularity; (2) composite Brainnetome Brain atlas (gyri) & Probabilistic Cerebellar atlas, which was of lowest granularity; and (3) AAL atlas (Tzourio-Mazoyer et al., 2002), which was of medium granularity, were used. To get a lobe level understanding of the links, they were mapped to Brainnetome brain atlas lobes. Also, to find patterns in the connectivity of multiple brain networks such as Visual, Somatomotor, DMN, Dorsal Attention, Ventral Attention, Limbic and Control, the links were mapped to Schaefer brain atlas (Schaefer et al., 2017). The results for an additional mapping on the composite Harvard Oxford atlas and Probabilistic Cerebellar atlas are presented in the Appendix A.2. The following mapping rules were followed. For each atlas, the coordinates that lie outside the atlas were assigned a region from the atlas by finding the nearest atlas region in 6 voxel cube centered at that coordinate (that corresponded to 12mm cube for AAL Atlas and 6mm cube for other atlases). AAL atlas contains the cerebrum as well as cerebellar regions, but Brainnetome and Harvard Oxford atlas are just cerebrum atlases. So they must be combined with a cerebellar atlas which may lead to inconsistencies at the places of overlap of the cerebrum and Cerebellar atlases. To assign a voxel, belonging to the overlap, to the ROI of one of the atlases, the ROI name given in the publication from which link was extracted was checked if it belonged to cerebral or cerebellar cortex. If the study has mentioned the coordinates to lie in the cerebral cortex, the ROI from cerebral atlas was assigned, and if the study mentions that the region is a cerebellar region, then ROI from cerebellar atlas was assigned. In some of the atlases, the hemispheric assignment of the coordinates was found to be inconsistent. For example, (1, -57, 30) is left precuneus according to AAL atlas even though the x-coordinate is positive. According to Harvard Oxford atlas, it is right precuneus. Consequently, some studies have reported ambiguous hemispheres. For example, Fisherman et al. in their study (Fishman et al., 2015) have reported a positive x (+9) coordinate for left precuneus. Burrows et al. (Burrows et al., 2016) reported left superior frontal gyrus with MNI coordinate (2, 58, 40). As the x-coordinate is positive, it is expected to be in the right hemisphere in Harvard Oxford atlas and Talairach-Tournoux atlas. But it is in left according to AAL atlas as well as CA_ML_18_MNIA atlas (in AFNI). Therefore for this review, the hemispheric information was assigned explicitly with a fixed rule. The coordinate is assigned to the left hemisphere if x-coordinate is negative and right hemisphere if x-coordinate is positive. In case a seed or ROI is expected to lie in both the hemispheres, then that region is assigned the hemisphere information - ‘center’ denoted by ‘C’. It is because the representative time course of an ROI is typically the mean of the time courses all the voxels lying in that ROI, so the mean represents both the left and right hemispheres. For example, a seed region of a 6mm radius is expected to lie in ‘center’ if the absolute value of x-coordinate is less than 6mm.

The reported peak coordinates (of the cluster of voxels significantly differently connected to seed in ASD as compared to TD), lie close to mid-line i.e. whose x-coordinate’s absolute value is less than 6mm, is duplicated with x-coordinate as 2 (along with the relevant sign) in the contra-lateral hemisphere with its y and z coordinates kept the same. A cluster reported to be differently connected to the seed is expected to span both hemispheres if the reported x-coordinate is less than 6mm. This is a heuristic based on the fact that majority of the studies take the seed to be a 6mm sphere. If in the seed-to-voxel based analysis, the x-coordinate of the target cluster specifying altered connectivity is given to be zero, then a duplicate link is introduced and x-coordinate in both the links (original and duplicate) modified to be -2 and +2 respectively. For example, anterior cingulate gyrus (ACC) cluster reported in the study by Verly et al. had MNI coordinates (0, 26, 28) (Verly et al., 2014). We introduced left ACC with coordinates (-2, 26, 28) and right ACC with coordinates (2, 26, 28). In ROI-to-ROI based studies, the ROI’s hemisphere is said to be ‘C’ if the ROI spans both the hemispheres. For example in study (Fishman et al., 2014) the ROI is constructed using a 6 mm sphere with the x-coordinate of less than 6 mm, then it is expected to cross the mid-line. For the studies such as the one by Linke et al. (Linke et al., 2017b) who defined the ROIs using an atlas parcellation, was assigned the hemisphere to which the parcellation belongs. In a study by Abbot et al. (Abbott et al., 2016) it was mentioned that the bilateral regions of the dorsal medial cingulate cortex were found to be differently connected to the seed. As only the left hemisphere coordinate was given, the contralateral ROI was set to be the same as the left ROI coordinate, but, with the sign of x-Coordinate negated. This strategy of assigning hemispheres can change if the author has explicitly mentioned that the seed region does not span the contra-lateral hemisphere or has mentioned about all the regions spanned by the cluster (about if they span the contra-lateral hemispheric region).

If any study has reported the coordinates in decimal form, for example, raw coordinates from FreeSurfer (Fischl, 2012) or center of mass, etc., the value after decimal was trimmed off. One of the studies by Padmanabhan et al. (Padmanabhan et al., 2013) reported the seed coordinates in MNI space in LAS orientation whereas the center of gravity coordinates of the target cluster in LPS. So the coordinates were transformed to RAS orientation by negating the x as well as y coordinates.

AAL atlas, Probabilistic Cerebellar atlas, and Harvard Oxford atlas regions have the hemispheric information associated with the regions. To follow the above defined rule of assigning hemispheres, the regions were renamed with their hemispheric information removed. For example, ‘Precuneus_L’ in AAL atlas was renamed to ‘Precuneus’, and the new hemispheric information (based on the above rules) was added. The names in the figures and the tables follow this new naming scheme.

Once the MNI coordinates were mapped back to atlas regions, the region and the hemisphere names were used to find the replications, consistencies, and inconsistencies in the literature. Recall from the section 2.4, a link is said to be consistent if that link reports the same connectivity across all the studies to which it belongs. A link is termed inconsistent when it states different connectivities across at least two studies to which it belongs. If there are inconsistent links between region R_1_ and region R_2_, the measures DR_1_ and DR_2_ (see Table 4) represent the smallest distance (in mm) between the endpoints of the two inconsistent links in the regions R_1_ and R_2_ respectively. For the inconsistent links between the regions R_1_ and R_2_, large values of DR_1_ and DR_2_ would mean that this large distance between the links could be a potential reason of the inconsistency.

### 2.8. Data and Code Availability Statement

The data extracted from the studies, and the code used to process it and derive conclusions are made available in the public domain. The data is available in the form of spreadsheet and the code is available on GitHub. See Appendix A.3 for the URLs.

## 3. Results

### 3.1. Characteristics of Included Studies

Table 1 shows the aggregate demographics and the main brain regions studied by the publications included in this review. A majority of 29 out of 44 studies had an average age between 10 and 20 years. Figure 3(B) shows the gender distribution of the studies reviewed. Majority of the studies (29 out of 44) investigated both male and female participants. Fourteen studies investigated only males and one investigated only females. Even though a majority of studies involved both males and females in the investigation, the female to male ratio is quite small.

**Figure 3:**
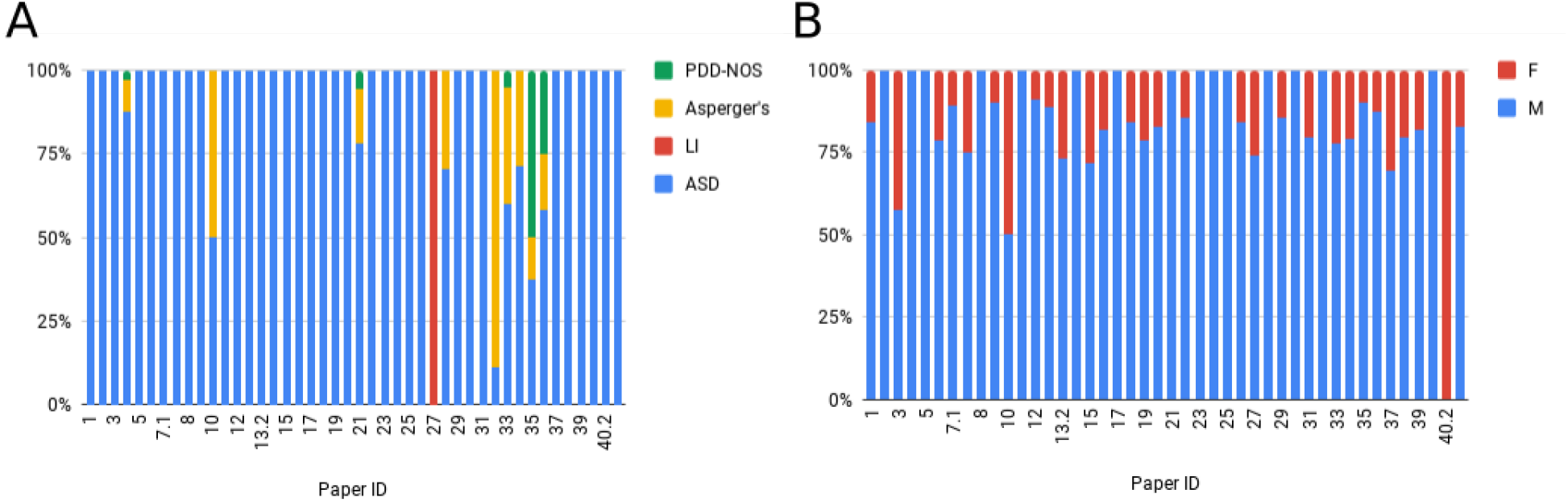
Per study distribution of (A) Percentage of ASD subtypes and language impaired participants; (B) Percentage of males and female participants. Abbreviations: PDD-NOS, Pervasive developmental disorder not otherwise specified; ASD, Autism spectrum disorder; LI, Language impaired; F, Female; M, Male.

Figure 4(A) and Figure 4(B) respectively show count of number of participants and its histogram. A majority of the studies investigated a small number of participants. Nineteen out of 44 studies investigated less than 50 participants, and 15 studies had 50 to 100 participants. Only one study crossed a 1000 participants mark.

**Figure 4:**
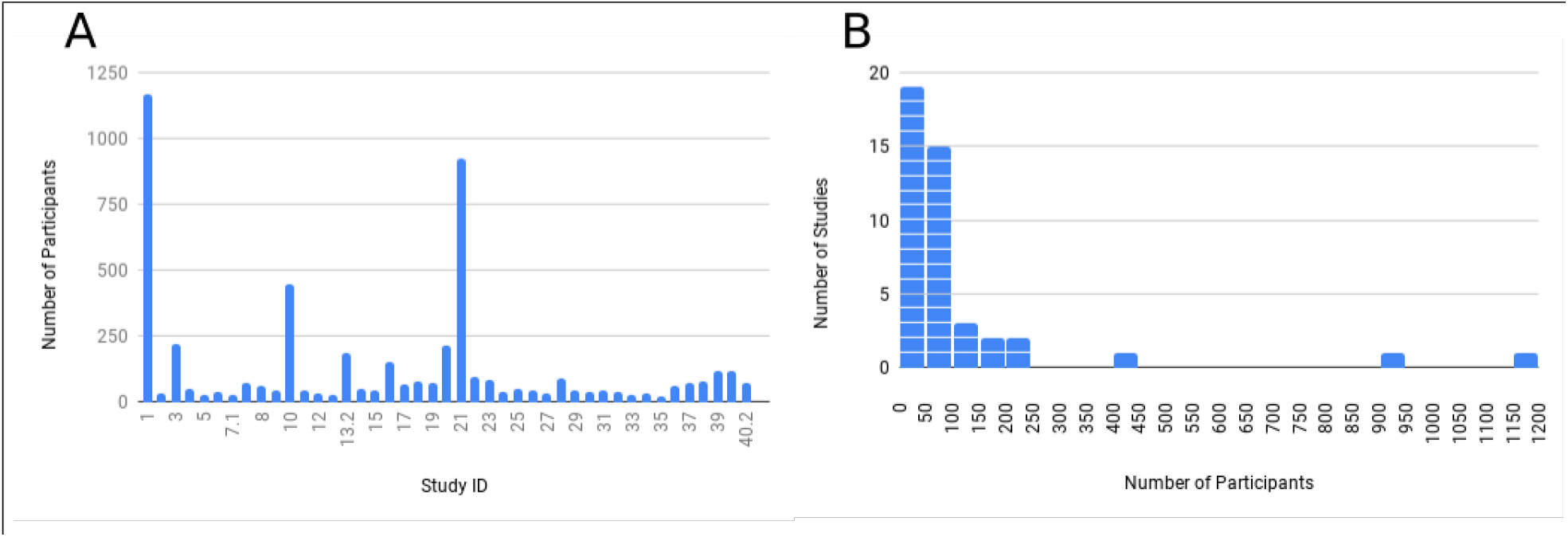
(A) Count of the number of participants per study. (B) Histogram showing the relationship between the number of participants and the number of studies that analyzed them.

Figure 3(A) shows the distribution of ASD subtypes constituting the population investigated in each of the studies. The studies which did not mention autism subtypes (autism, Aspergers’ syndrome and PDD-NOS) of the participants, have reported the participants to have ASD. Participants diagnosed with ASD according to DSM-IV comprised of autism, Asperger’s syndrome and PDD-NOS. One study looked at autistic participants with language impairment.

Clinical diagnoses of ASD was according to DSM-IV in 28 studies. Eight studies adopted DSM-V. One study had a mix of participants, some diagnosed according to DSM-IV and others according to DSM-V. Seven studies did not specify the DSM version employed for the diagnosis of ASD.

Out of 44 studies, 14 studies utilized ABIDE-I data. Out of those 14, one also used ABIDE-II data, and another one had some participants from ABIDE-I and rest were independently scanned. The remaining 30 studies collected their data. The total number of subjects was calculated as the sum of the number of subjects for each study that collected their data and the number of subjects of the study that employed the maximum number of ABIDE’s subjects. Majority of the studies (19 out of 44) had the participants with their eyes open during the scan. There is an almost equal proportion of studies who had participants with their eyes closed (10) as well as both open and closed (9). Three studies had their participants sleeping - one in natural sleep and two in sedated sleep.

To tackle physiological noise, the majority of publications regressed out motion parameters (38/41), CSF (36/41) and WM (35/41) as nuisance covariates. Other nuisance covariates included to remove physiological noise were: global correlation (GOR) that was regressed out by two studies, global brain signal (GS) that was removed by 17 studies and cardiac and respiratory measurements (CR) which was regressed out by five studies. Refer to Table 2 for more details. All of the studies used a 3 Tesla magnetic field strength scanner.

Three publications did both ROI-to-ROI (called ROI-based here) as well as seed-to-voxels (called seed-based here) based analysis (represented as RB and SB in the Table 1). Six publications performed just ROI-to-ROI based analysis out of which one (Cheng et al., 2015) first did voxels-to-voxels based analysis to select relevant voxels and then performed ROI-to-ROI based analysis after mapping only those voxels to anatomical regions by using an atlas hence we call it an ROI-to-ROI based analysis. The remaining 32 publications did the seed-to-voxels based analysis.

### 3.2. Summary of Brain Regions Studied

A total of 932 links were extracted out of 44 studies selected for review. Both the endpoints of each of the extracted links were mapped to AAL atlas (see section 2.7). Figure 5 shows the count of altered connectivity links associated with different brain regions of the AAL atlas. It is observed that the middle temporal region given by name Temporal_Mid connects to the highest number of altered connectivity links. The reason for this is that this region is very large. Looking at the other regions, we can see that the amygdala has the next highest reported altered connectivity links with the majority of links being underconnected. The DMN regions - precuneus, posterior cingulate cortex (PCC), anterior cingulate cortex (ACC); and the salience network’s (SN) region insula comes next.

**Figure 5:**
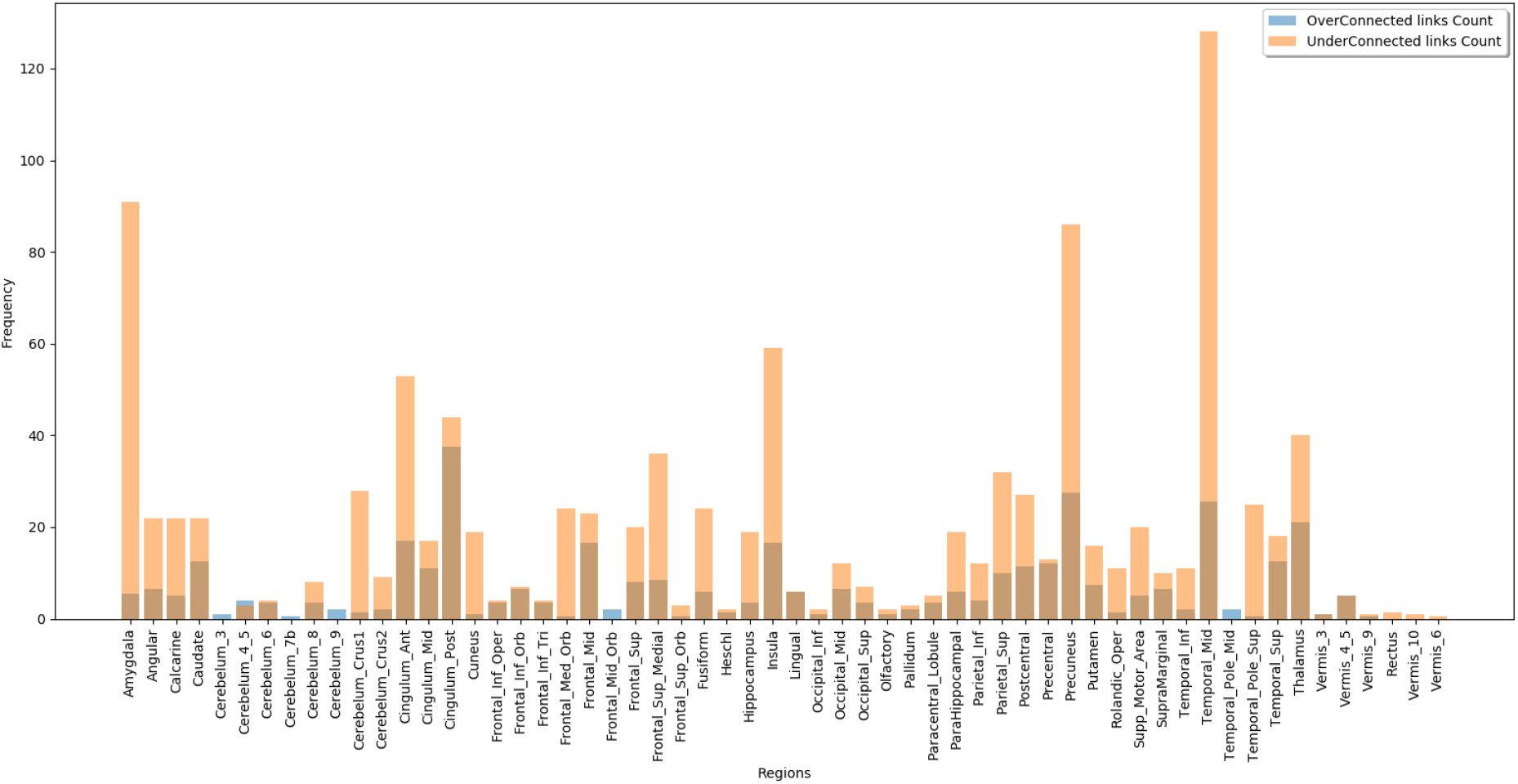
The figure shows the number of altered connectivity links associated with each of the AAL regions. The Blue/Dark color bars denote the count of overconnected links; Orange color bars represent the count of underconnected links.

The most reported regions of altered functional connectivity were in the Default mode network (DMN) including PCC, precuneus and medial prefrontal cortex (mPFC). Majority of the studies investigating DMN (Doyle-Thomas et al., 2015), (Eilam-Stock et al., 2014), (Monk et al., 2009), (Yerys et al., 2015), (Traynor et al., 2018) reported a mix of over and under connectivity. Quite a few studies (Burrows et al., 2016), (Jung et al., 2014), (Lynch et al., 2013), (Weng et al., 2010) reported just underconnectivity and one study (Lynch et al., 2013) reported just overconnectivity for DMN.

Salience Network and its subregions such as amygdala and insula were the next most studied region. With multiple studies (Eilam-Stock et al., 2014), (Shou et al., 2017), (Guo et al., 2016), (Shen et al., 2016), (Rausch et al., 2016) reporting underconnectivity and overconnectivity (Guo et al., 2016), (Green et al., 2016), (Ebisch et al., 2011). Some studies investigated multiple amygdaloid subregions as well and observed superficial (SF) nuclei and laterobasal (LB) nuclei to be under connected with the rest of the brain regions (Rausch et al., 2016), (Kleinhans et al., 2016). Overconnectivity for centromedial and superficial nuclei were also reported (Kleinhans et al., 2016).

Various other cortical regions involved in social cognitive processes such as the thalamus, pSTS, and TPJ were also investigated in the reviewed literature. Thalamus was reported to be majorly overconnected by multiple studies (Tomasi and Volkow, 2019), (Linke et al., 2017a), (Traynor et al., 2018). Instances of both under and over thalamocortical connectivity were also reported (Nair et al., 2015). Posterior STS (pSTS) was also reported to be just underconnected (Alaerts et al., 2015). Temporoparietal junction (TPJ), also considered to be involved in mentalizing and the Theory of Mind (TOM) was reported to be overconnected (Chien et al., 2015) as well as under connected (Hoffmann et al., 2016). For the putative involvement of basal ganglian structures in cognitive and affective function, Padmanabhan et al. analyzed the resting state functional connectivity of a subcortical brain structure called striatum and observed overconnectivity (Padmanabhan et al., 2013).

Delmonte et al. investigated the frontostriatal circuitry and observed increased functional connectivity between the frontal cortex’s regions and striatum’s regions in ASD individuals (Delmonte et al., 2013). Traynor et al. analyzed the cortico-basal ganglia regions and observed a predominant overconnectivity of basal gangalian regions - putamen and globus pallidus seeds with cortical sensory processing areas (Traynor et al., 2018). Overconnectivity of multiple caudal seeds, multiple putamen seeds as well as underconnectivity of one putamen seed was reported (Di Martino et al., 2011). As the autism prevalence in females is very low as compared to males, Alaerts et al. explored the functional brain connectivity differences in ASD males and females. Upon independent analysis of 2 groups - males and females, they observed predominant underconnectivity in ASD males as compared to TD males whereas overconnectivity in female ASD participants as compared to TD females (Alaerts et al., 2016).

### 3.3. Replication Results with AAL Atlas

Recall that, a total of 932 links were extracted out of 44 studies selected for review, out of which 927 got mapped to AAL atlas. These 927 non-unique links translated to 645 unique links. Out of the 645 unique links, 71 links were replicated, i.e., each of the 71 links was reported by at least two studies. These 71 unique replicated links corresponded to 49 consistent links (33 underconnected and 16 overconected). The consistent links constitute the AAFC which is depicted in graphical form in Figure 6. Out of 49 unique, consistent links, 38 links were found to be perfectly replicated. The remaining 11 links were replicated but not perfectly. Refer to the Table 3 for the complete list of replicated links along with information about which of them were perfectly replicated. Out of the total 645 unique links that resulted after mapping the 932 links to AAL atlas, 574 unique links were not replicated. That is, each of these unique links had only one study reporting it. Also, out of 71 unique replicated links, and 22 unique links were observed to be inconsistent.

**Figure 6:**
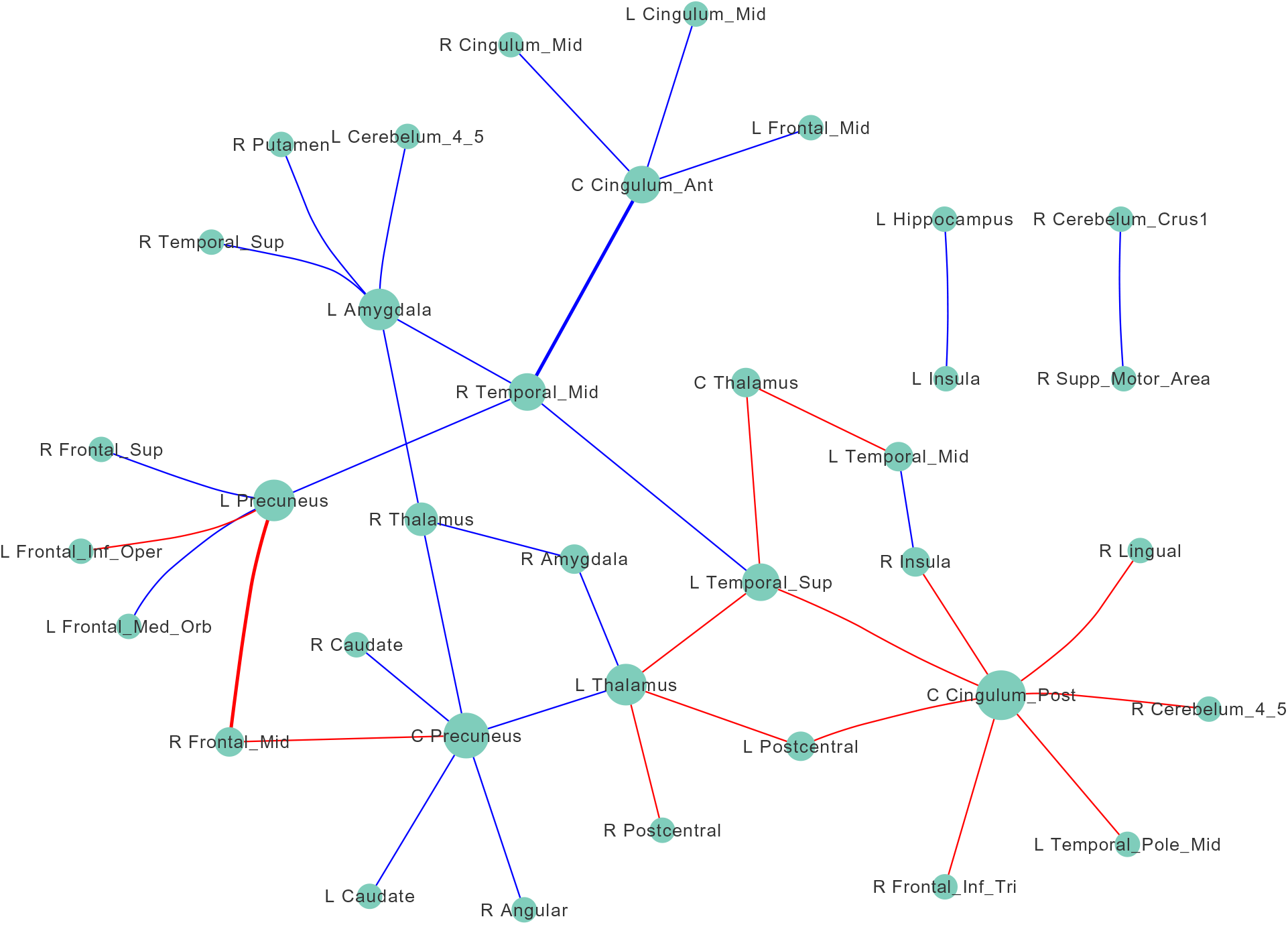
Autism Altered Functional Connectome (AAL Atlas): Blue edges represent underconnectivity (i.e., Autistic *<* TD) and red edges represent overconnectivity. Node size is proportional to the degree of the node. Edge width is proportional to the number of studies with which the link is consistent. Abbreviations: L, Left; R, Right; Sup, Superior; Orb, Orbital; Mid, Middle; Inf, Inferior; Oper, Operculum; Tri, Triangular; Supp, Supplementary; Ant, Anterior; Post, Posterior.

**Table 3.**
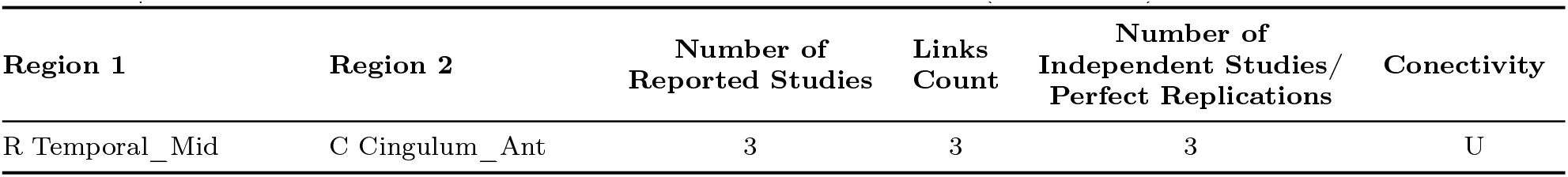
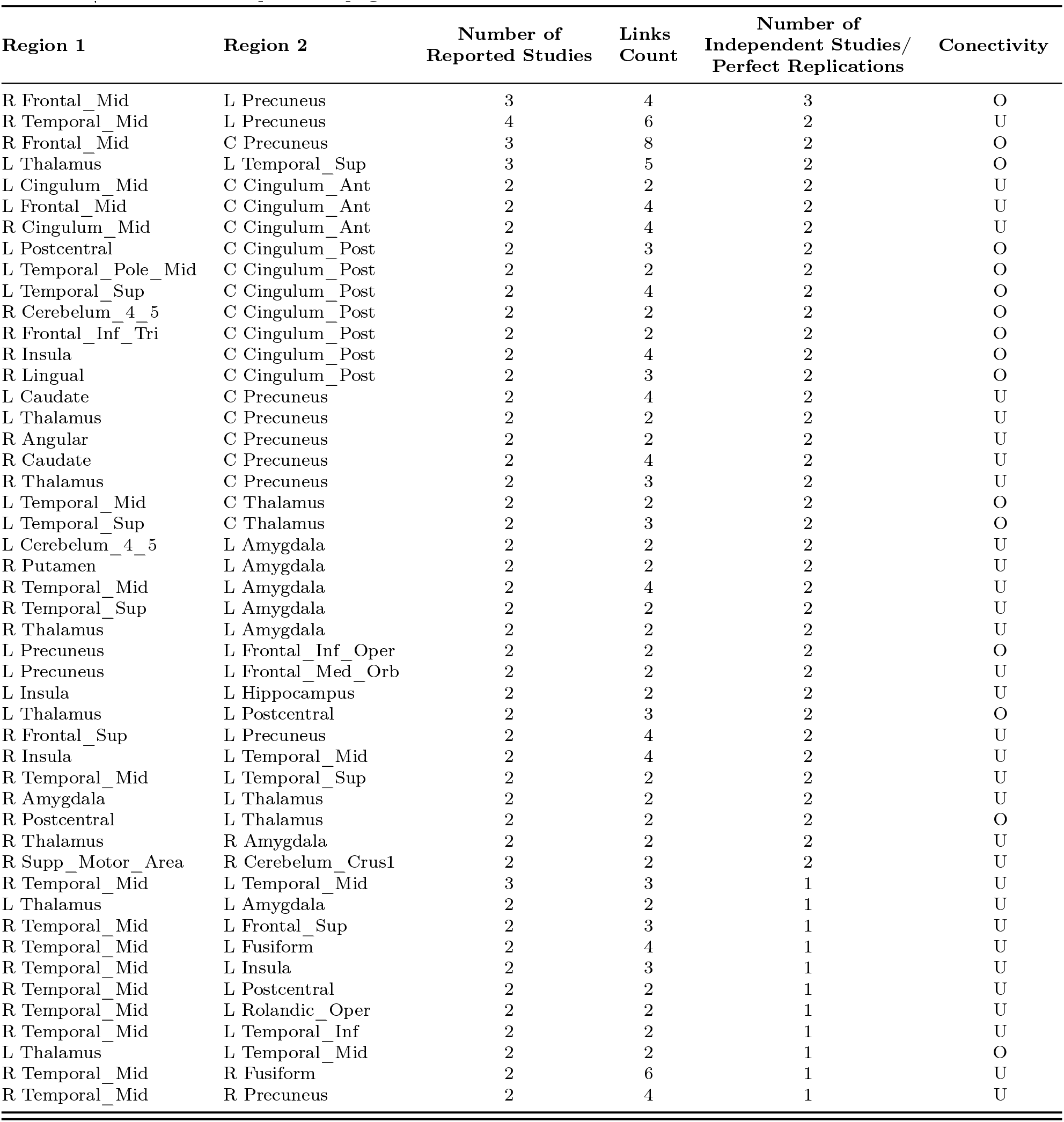
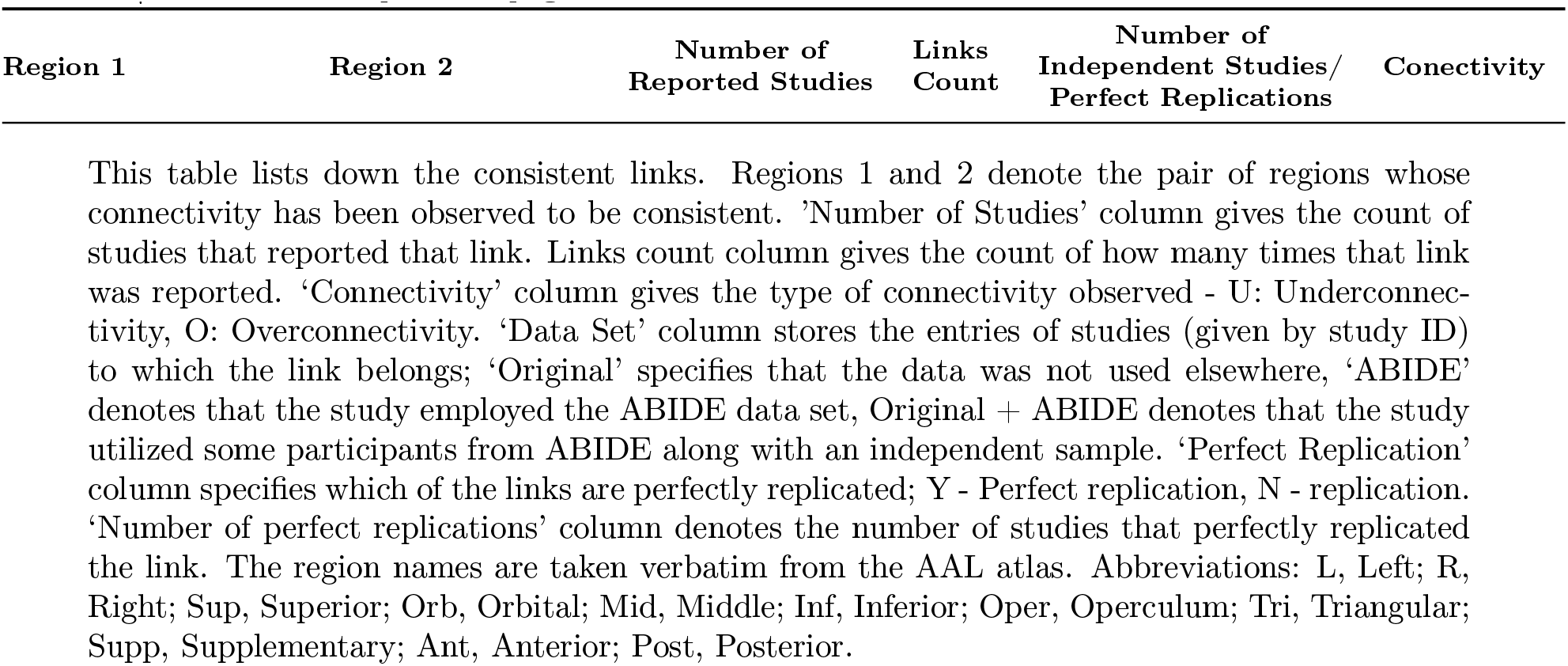
Consistent Autism Altered Functional Connectivity Links (AAL Atlas)

| Region 1 | Region 2 | Number of<br>Reported Studies | Links<br>Count | Number of<br>Independent Studies/<br>Perfect Replications | Conectivity |
| --- | --- | --- | --- | --- | --- |
| R Temporal_Mid | C Cingulum_Ant | 3 | 3 | 3 | U |

Table 3 | continued from previous page
| Region 1 | Region 2 | Number of<br>Reported Studies | Links<br>Count | Number of<br>Independent Studies/<br>Perfect Replications | Conectivity |
| --- | --- | --- | --- | --- | --- |
| R Frontal_Mid | L Precuneus | 3 | 4 | 3 | O |
| R Temporal_Mid | L Precuneus | 4 | 6 | 2 | U |
| R Frontal_Mid | C Precuneus | 3 | 8 | 2 | O |
| L Thalamus | L Temporal_Sup | 3 | 5 | 2 | O |
| L Cingulum_Mid | C Cingulum_Ant | 2 | 2 | 2 | U |
| L Frontal_Mid | C Cingulum_Ant | 2 | 4 | 2 | U |
| R Cingulum_Mid | C Cingulum_Ant | 2 | 4 | 2 | U |
| L Postcentral | C Cingulum_Post | 2 | 3 | 2 | O |
| L Temporal_Pole_Mid | C Cingulum_Post | 2 | 2 | 2 | O |
| L Temporal_Sup | C Cingulum_Post | 2 | 4 | 2 | O |
| R Cerebelum_4_5 | C Cingulum_Post | 2 | 2 | 2 | O |
| R Frontal_Inf_Tri | C Cingulum_Post | 2 | 2 | 2 | O |
| R Insula | C Cingulum_Post | 2 | 4 | 2 | O |
| R Lingual | C Cingulum_Post | 2 | 3 | 2 | O |
| L Caudate | C Precuneus | 2 | 4 | 2 | U |
| L Thalamus | C Precuneus | 2 | 2 | 2 | U |
| R Angular | C Precuneus | 2 | 2 | 2 | U |
| R Caudate | C Precuneus | 2 | 4 | 2 | U |
| R Thalamus | C Precuneus | 2 | 3 | 2 | U |
| L Temporal_Mid | C Thalamus | 2 | 2 | 2 | O |
| L Temporal_Sup | C Thalamus | 2 | 3 | 2 | O |
| L Cerebelum_4_5 | L Amygdala | 2 | 2 | 2 | U |
| R Putamen | L Amygdala | 2 | 2 | 2 | U |
| R Temporal_Mid | L Amygdala | 2 | 4 | 2 | U |
| R Temporal_Sup | L Amygdala | 2 | 2 | 2 | U |
| R Thalamus | L Amygdala | 2 | 2 | 2 | U |
| L Precuneus | L Frontal_Inf_Oper | 2 | 2 | 2 | O |
| L Precuneus | L Frontal_Med_Orb | 2 | 2 | 2 | U |
| L Insula | L Hippocampus | 2 | 2 | 2 | U |
| L Thalamus | L Postcentral | 2 | 3 | 2 | O |
| R Frontal_Sup | L Precuneus | 2 | 4 | 2 | U |
| R Insula | L Temporal_Mid | 2 | 4 | 2 | U |
| R Temporal_Mid | L Temporal_Sup | 2 | 2 | 2 | U |
| R Amygdala | L Thalamus | 2 | 2 | 2 | U |
| R Postcentral | L Thalamus | 2 | 2 | 2 | O |
| R Thalamus | R Amygdala | 2 | 2 | 2 | U |
| R Supp_Motor_Area | R Cerebelum_Crus1 | 2 | 2 | 2 | U |
| R Temporal_Mid | L Temporal_Mid | 3 | 3 | 1 | U |
| L Thalamus | L Amygdala | 2 | 2 | 1 | U |
| R Temporal_Mid | L Frontal_Sup | 2 | 3 | 1 | U |
| R Temporal_Mid | L Fusiform | 2 | 4 | 1 | U |
| R Temporal_Mid | L Insula | 2 | 3 | 1 | U |
| R Temporal_Mid | L Postcentral | 2 | 2 | 1 | U |
| R Temporal_Mid | L Rolandic_Oper | 2 | 2 | 1 | U |
| R Temporal_Mid | L Temporal_Inf | 2 | 2 | 1 | U |
| L Thalamus | L Temporal_Mid | 2 | 2 | 1 | O |
| R Temporal_Mid | R Fusiform | 2 | 6 | 1 | U |
| R Temporal_Mid | R Precuneus | 2 | 4 | 1 | U |

**Table 3 | continued from previous page**
| **Region 1** | **Region 2** | **Number of**<br>**Reported Studies** | **Links**<br>**Count** | **Number of**<br>**Independent Studies/**<br>**Perfect Replications** | **Conectivity** |
| --- | --- | --- | --- | --- | --- |

This table lists down the consistent links. Regions 1 and 2 denote the pair of regions whose connectivity has been observed to be consistent. 'Number of Studies' column gives the count of studies that reported that link. Links count column gives the count of how many times that link was reported. 'Connectivity' column gives the type of connectivity observed - U: Underconnectivity, O: Overconnectivity. 'Data Set' column stores the entries of studies (given by study ID) to which the link belongs; 'Original' specifies that the data was not used elsewhere, 'ABIDE' denotes that the study employed the ABIDE data set, Original + ABIDE denotes that the study utilized some participants from ABIDE along with an independent sample. 'Perfect Replication' column specifies which of the links are perfectly replicated; Y - Perfect replication, N - replication. 'Number of perfect replications' column denotes the number of studies that perfectly replicated the link. The region names are taken verbatim from the AAL atlas. Abbreviations: L, Left; R, Right; Sup, Superior; Orb, Orbital; Mid, Middle; Inf, Inferior; Oper, Operculum; Tri, Triangular; Supp, Supplementary; Ant, Anterior; Post, Posterior.

**Table 4.**
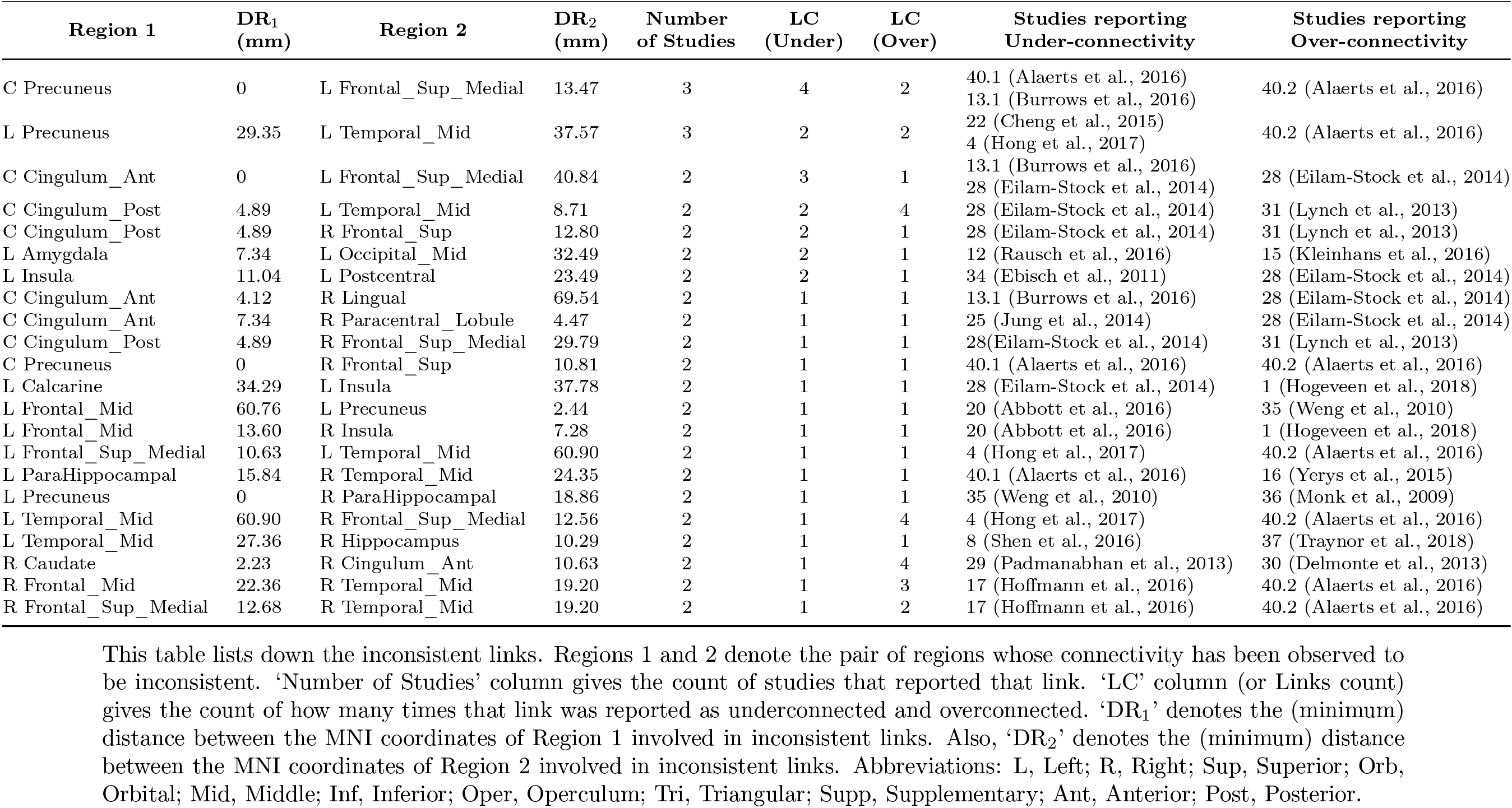
Inconsistent Autism Altered Functional Connectivity Links (AAL Atlas)

Maximum consistency was seen in the study by Alaerts et al. (study ID 21) (Alaerts et al., 2015). This study reported nine consistent links and no inconsistent link. Another study observed to be entirely consistent was a study by Guo et al. (study ID 7) (Guo et al., 2016) which reported seven consistent links after mapping to AAL atlas.

#### 3.3.1. Consistent Replicated Links

Though the Table 3 shows that right middle temporal gyrus (R Temporal_Mid) has the maximum degree, i.e., it is common to the maximum number of consistently replicated links in the literature, upon considering the perfectly replicated links depicted in the Figure 6, it is observed that PCC has the highest degree .

The perfectly consistently replicated links that replicated across the highest number of studies were - link between left precuneus and right middle frontal gyrus (MFG) that presented overconnectivity; and link between central ACC and right middle temporal gyrus (MTG) presented consistent underconnectivity. Each of the links were replicated by 3 studies. Underconnectivity between Amygdala and Thalamus was observed which involved both the hemispheres. Amygdala was reported to be consistently underconnected.

Several regions of DMN were observed to be consistently differently connected. Posterior cingulate gyrus (Cingulum_post) was observed to be overconnected in autistic subjects. After mapping of links to the Harvard Oxford atlas, consistent underconnectivity of PCC is observed only with Precuneus and paracingulate gyrus which overlaps with mPFC. Precuneus was observed to be majorly underconnected comprising ten underconnected links and only three overconnected links.

#### 3.3.2. Inconsistent Replicated Links

Table 4 lists all the 22 inconsistent links along with the study ID and reference of studies to which a link belongs. One possible reason for the inconsistencies could be the gender differences. Alaerts et al. in one of their studies analyzed only female participants (Alaerts et al., 2016) (study ID 40.2). This study was inconsistent with five studies which had a majority of males. It resulted in seven inconsistent links - central precuneus and left medial superior frontal cortex, left precuneus and left middle temporal gyrus, central ACC and left medial superior frontal cortex, central PCC and left middle temporal gyrus and right superior frontal gyrus, left amygdala and left middle occipital gyrus and between left insula and left postcentral gyrus. Out of these five studies, one study was from the same publication (Alaerts et al., 2016) (study ID 40.1) that analyzed all male participants. Kleinhans and colleagues in their study (study ID 15) (Kleinhans et al., 2016) found the link between the left amygdala and left middle occipital gyrus to be overconnected whereas it was observed underconnected in the study by Rausch et al. (Rausch et al., 2016) (study ID 12). Out of total ASD participants, the former had 56.25 % females whereas the later had just 5.2% females.

Age-related factors may also contribute to the conflicting connectivity findings between regions (Kana et al., 2014). Connectivity links between central PCC and left middle temporal gyrus, and right superior frontal gyrus and right medial superior frontal gyrus were observed to be inconsistent between the studies by Lynch et al. (Lynch et al., 2013) and Eilam-Stock et al. (Eilam-Stock et al., 2014). Participants in the study by Lynch et al. (study ID 31) were children with a mean age of ASD and TD being 9.88 and 9.96 years respectively. This study reported overconnectivity for the above-mentioned links. Whereas in the study by Eilam-Stock et al. (study ID 28) participants were adults with the mean age of ASD and TD being 27.1 and 26.1 years respectively. They reported under-connectivity among these links. The link between left middle temporal gyrus and right hippocampus was found to be inconsistent across the two studies, one by Shen et al. (Shen et al., 2016) and other by Traynor and colleagues (Traynor et al., 2018) who analyzed different age groups. Shen et al. (study ID 8) analyzed young sleeping children with mean age of ASD and TD being 3.6 and 3.5 respectively and reported overconnectivity whereas Traynor and colleagues (study ID 37) examined awake adolescents with a mean age of ASD and TD being 15 and 16 respectively and reported underconnectivity for the specified link.

### 3.4. Replication Results with Brainnetome & Cerebellar Atlases

To see the impact of atlas granularity on replication results, the MNI coordinates were mapped onto a composite Brainnetome atlas and Probabilistic Cerebellar atlas, and replication results are reported at two granularities. The finer granularity results employ the region names of the Brainnetome atlas, which gives rise to a total of 274 regions spanning the cerebrum as well as the cerebellum. This resolution is finer than the AAL atlas which comprises 116 regions. This mapping results into just 20 consistencies comprising of 14 unique underconnected links and six unique overconnected links which can be seen in the Table 5. The consistent connectome can be seen in the Figure 7. Also, this mapping resulted in just three inconsistent links which can be seen in Table 6. Each of the consistent links was replicated by exactly two studies.

**Figure 7:**
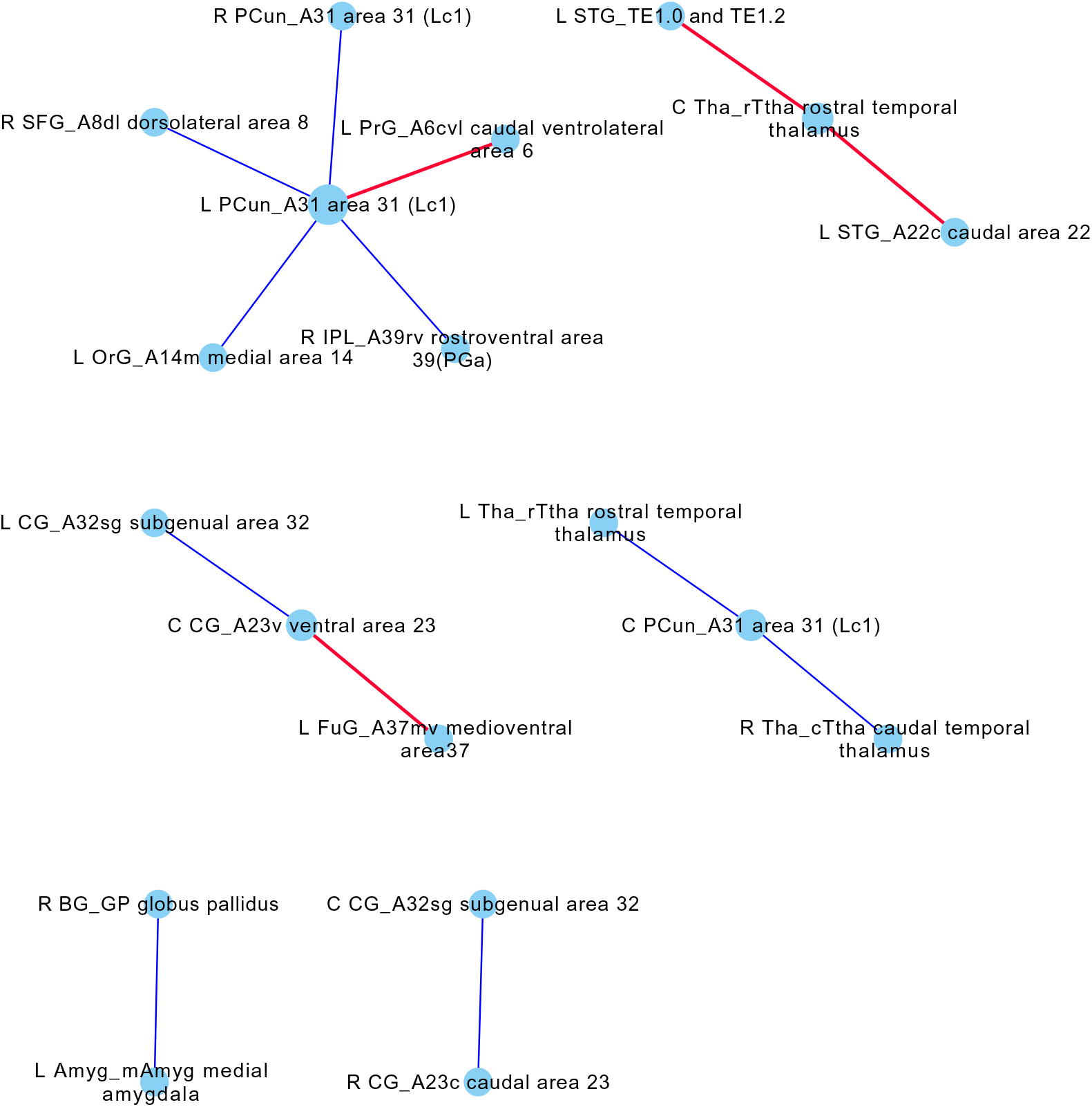
Autism Altered Functional Connectome (Brainnetome (Regions) & Probabilistic Cerebellar Atlases): Blue edges represent under-connectivity (i.e. Autistic *<* TD) and red edges represent over-connectivity. Node size is proportional to the degree of the node. Edge width is proportional to the number of studies with which the link is consistent. Abbreviations: L, Left; R, Right; C, Center; Amyg, Amygdala; BG, Basal Ganglia; CG, Cingulate Gyrus; FuG, Fusiform Gyrus; Hipp, Hippocampus; IFG, Inferior Frontal Gyrus; INS, Insular Gyrus; IPL, Inferior Parietal Lobule; ITG, Inferior Temporal Gyrus; LOcC, lateral Occipital Cortex; MFG, Middle Frontal Gyrus; MTG, Middle Temporal Gyrus; MVOcC, MedioVentral Occipital Cortex; OrG, Orbital Gyrus; PCL, Paracentral Lobule; Pcun, Pre-cuneus; PhG, Parahippocampal Gyrus; PoG, Postcentral Gyrus; PrG, Precentral Gyrus; pSTS, posterior Superior Temporal Sulcus; SFG, Superior Frontal Gyrus; SPL, Superior Parietal Lobule; STG, Superior Temporal Gyrus; Tha, Thalamus.

**Table 5.** Consistent Autism Altered Functional Connectivity Links (Brainnetome (Regions) & Cerebellar Atlases)

| Region 1 | Region 2 | Number of Reported Studies | Links Count | Number of Independent Studies/<br>Perfect Replications | Connectivity |
| --- | --- | --- | --- | --- | --- |
| L CG_A32sg, subgenual area 32 | C CG_A23v, ventral area 23 | 2 | 2 | 2 | U |
| L FuG_A37mv, medioventral area37 | C CG_A23v, ventral area 23 | 2 | 2 | 2 | O |
| R CG_A23c, caudal area 23 | C CG_A32sg, subgenual area 32 | 2 | 4 | 2 | U |
| L Tha_rTtha, rostral temporal thalamus | C PCun_A31, area 31 (Lcl) | 2 | 2 | 2 | U |
| R Tha_cTtha, caudal temporal thalamus | C PCun_A31, area 31 (Lcl) | 2 | 2 | 2 | U |
| L STG_A22c, caudal area 22 | C Tha_rTtha, rostral temporal thalamus | 2 | 2 | 2 | O |
| L STG_TE1.0 and TE1.2 | C Tha_rTtha, rostral temporal thalamus | 2 | 2 | 2 | O |
| R BG_GP, globus pallidus | L Amyg_mAmyg, medial amygdala | 2 | 2 | 2 | U |
| L PCun_A31, area 31 (Lcl) | L OrG_A14m, medial area 14 | 2 | 2 | 2 | U |
| L PrG_A6cvl, caudal ventrolateral area 6 | L PCun_A31, area 31 (Lcl) | 2 | 2 | 2 | O |
| R IPL_A39rv, rostroventral area 39(PGa) | L PCun_A31, area 31 (Lcl) | 2 | 2 | 2 | U |
| R PCun_A31, area 31 (Lcl) | L PCun_A31, area 31 (Lcl) | 2 | 2 | 2 | U |
| R SFG_A8dl, dorsolateral area 8 | L PCun_A31, area 31 (Lcl) | 2 | 4 | 2 | U |
| R MFG_A9/46v, ventral area 9/46 | C PCun_A31, area 31 (Lcl) | 2 | 2 | 1 | O |
| R MTG_A37dl, dorsolateral area37 | L FuG_A20rv, rostroventral area 20 | 2 | 4 | 1 | U |
| R MTG_A37dl, dorsolateral area37 | L FuG_A37lv, lateroventral area37 | 2 | 2 | 1 | U |
| R MTG_A37dl, dorsolateral area37 | L INS_dlg, dorsal granular insula | 2 | 2 | 1 | U |
| L Tha_Otha, occipital thalamus | L STG_A22c, caudal area 22 | 2 | 2 | 1 | O |
| R MTG_A37dl, dorsolateral area37 | R FuG_A37lv, lateroventral area37 | 2 | 3 | 1 | U |
| R MTG_A37dl, dorsolateral area37 | R FuG_A37mv, medioventral area37 | 2 | 4 | 1 | U |

This table lists the links that were observed to be consistent after mapping the links to Brainnetome (regions) & cerebellar atlas.
Regions 1 and 2 denote the pair of regions whose connectivity has been observed to be consistent. 'Number of Studies' column gives the count of studies that reported that link. 'Number of Links' column gives the number of instances a particular link was reported.
Conn 'column' tells if the link was reported as underconnected or overconnected. Abbreviations: Conn, Connectivity; L, Left; R, Right; C, Center; Amyg, Amygdala; BG, Basal Ganglia; CG, Cingulate Gyrus; FuG, Fusiform Gyrus; Hipp, Hippocampus; IFG, Inferior Frontal Gyrus; INS, Insular Gyrus; IPL, Inferior Parietal Lobule; ITG, Inferior Temporal Gyrus; LOcC, lateral Occipital Cortex; MFG, Middle Frontal Gyrus; MTG, Middle Temporal Gyrus; MVOcC, MedioVentral Occipital Cortex; OrG, Orbital Gyrus; PCL, Paracentral Lobule; PCun, Precuneus; PhG, Parahippocampal Gyrus; PoG, Postcentral Gyrus; PrG, Precentral Gyrus; pSTS, posterior Superior Temporal Sulcus; SFG, Superior Frontal Gyrus; SPL, Superior Parietal Lobule; STG, Superior Temporal Gyrus; Tha, Thalamus.

**Table 6.**
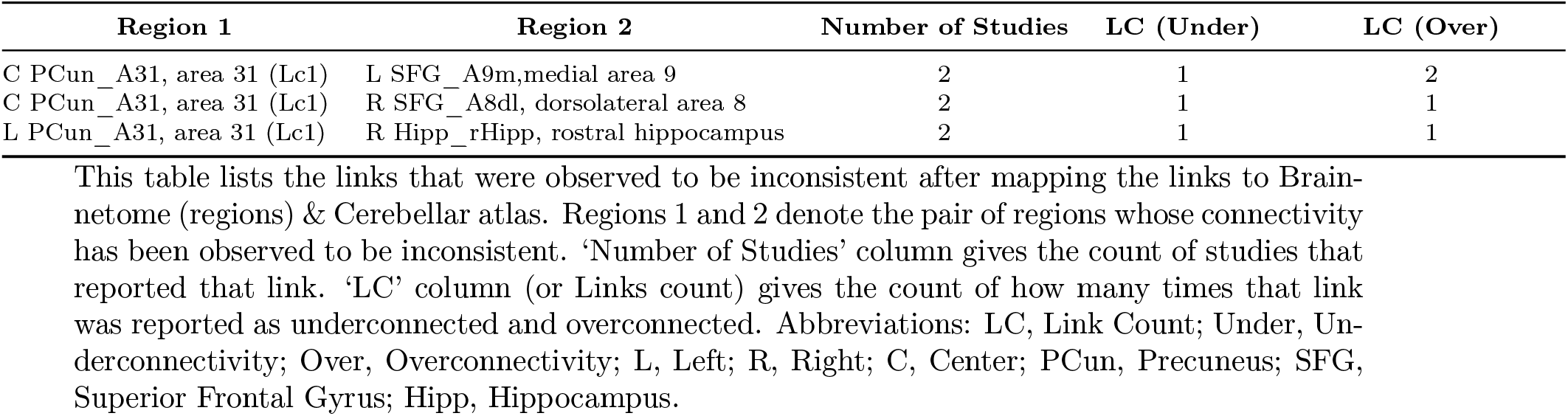
Inconsistent Autism Altered Functional Connectivity Links (Brainnetome (Regions) & Cerebellar Atlases)

| Region 1 | Region 2 | Number of Studies | LC (Under) | LC (Over) |
| --- | --- | --- | --- | --- |
| C PCun_A31, area 31 (Lc1) | L SFG_A9m,medial area 9 | 2 | 1 | 2 |
| C PCun_A31, area 31 (Lc1) | R SFG_A8dl, dorsolateral area 8 | 2 | 1 | 1 |
| L PCun_A31, area 31 (Lc1) | R Hipp_rHipp, rostral hippocampus | 2 | 1 | 1 |

This table lists the links that were observed to be inconsistent after mapping the links to Brainnetome (regions) & Cerebellar atlas. Regions 1 and 2 denote the pair of regions whose connectivity has been observed to be inconsistent. ‘Number of Studies’ column gives the count of studies that reported that link. ‘LC’ column (or Links count) gives the count of how many times that link was reported as underconnected and overconnected. Abbreviations: LC, Link Count; Under, Underconnectivity; Over, Overconnectivity; L, Left; R, Right; C, Center; PCun, Precuneus; SFG, Superior Frontal Gyrus; Hipp, Hippocampus.

The coarser granularity mapping uses Brainnetome gyri descriptions (in addition to cerebellum region names) which had a total of 76 ROIs. This mapping results in 51 unique, consistent links out of which 34 were underconnected and 17 were overconnected. The consistent AAFC for this mapping is shown graphically in the Figure 8 along with accompanying Table 7. Interestingly this mapping resulted in 50 unique inconsistent links which can be seen in the Table 8. It would be inappropriate to draw any meaningful conclusions from these inconsistencies because, at this level of granularity, the regions become too big and may have multiple functions with different functional connectivities. For example, the cingulate gyrus in the Brainnetome atlas has a volume of 46125 mm^3^ and spans two functionally different regions - anterior cingulate cortex (ACC) and posterior cingulate cortex (PCC). ACC is known to subserve executive functions whereas PCC to subserve evaluative functions (Vogt et al., 1992). And the current review, using AAL atlas, observed ACC to be consistently underconnected whereas PCC to be consistently overconnected to the other brain regions.

**Figure 8:**
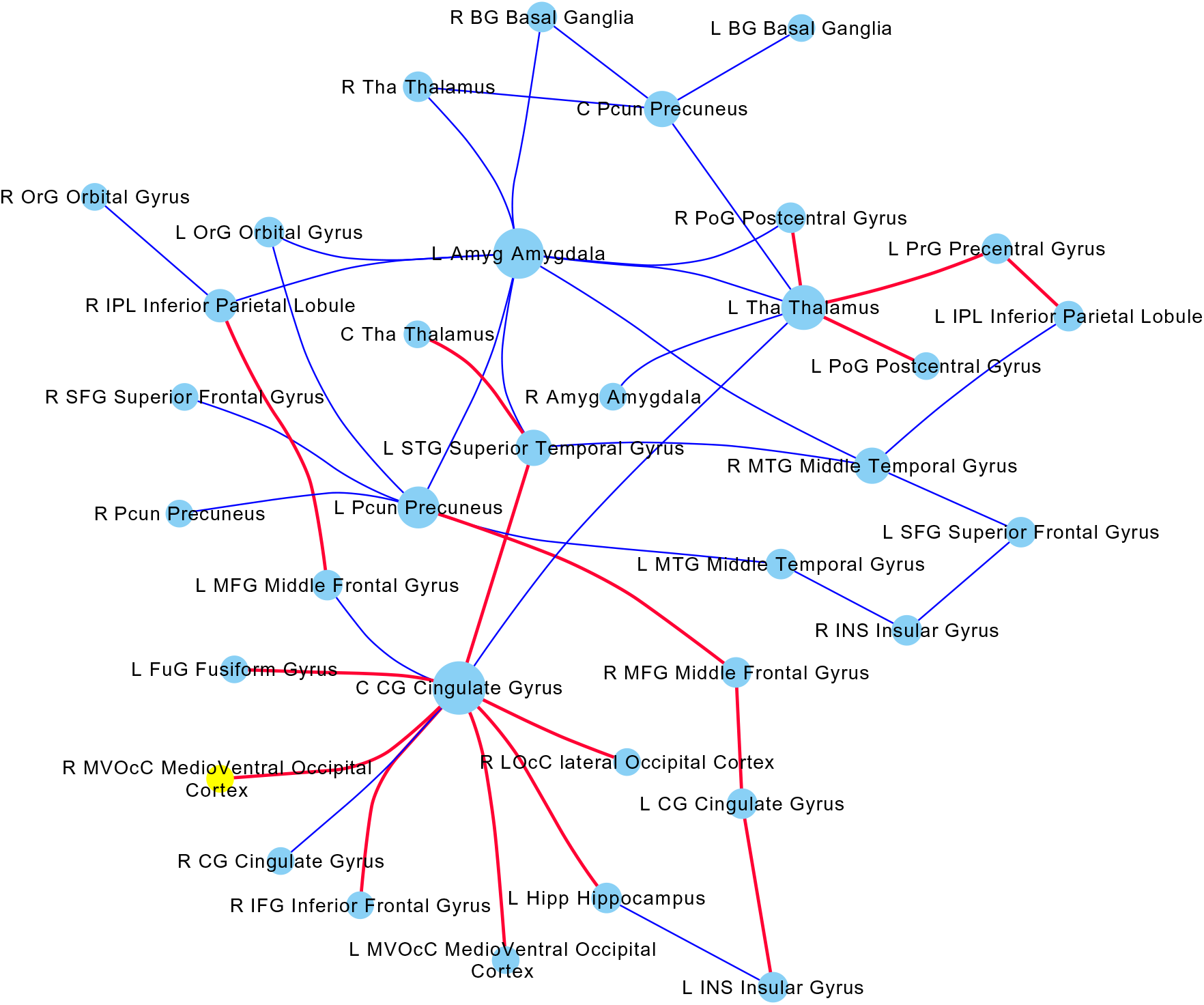
Autism Altered Functional Connectome (Brainnetome (Gyri) & Probabilistic Cerebellar Atlases): Blue edges represent under-connectivity (i.e. Autistic *<* TD) and red edges represent over-connectivity. Node size is proportional to the degree of the node. Edge width is proportional to the number of studies with which the link is consistent.

**Table 7.**
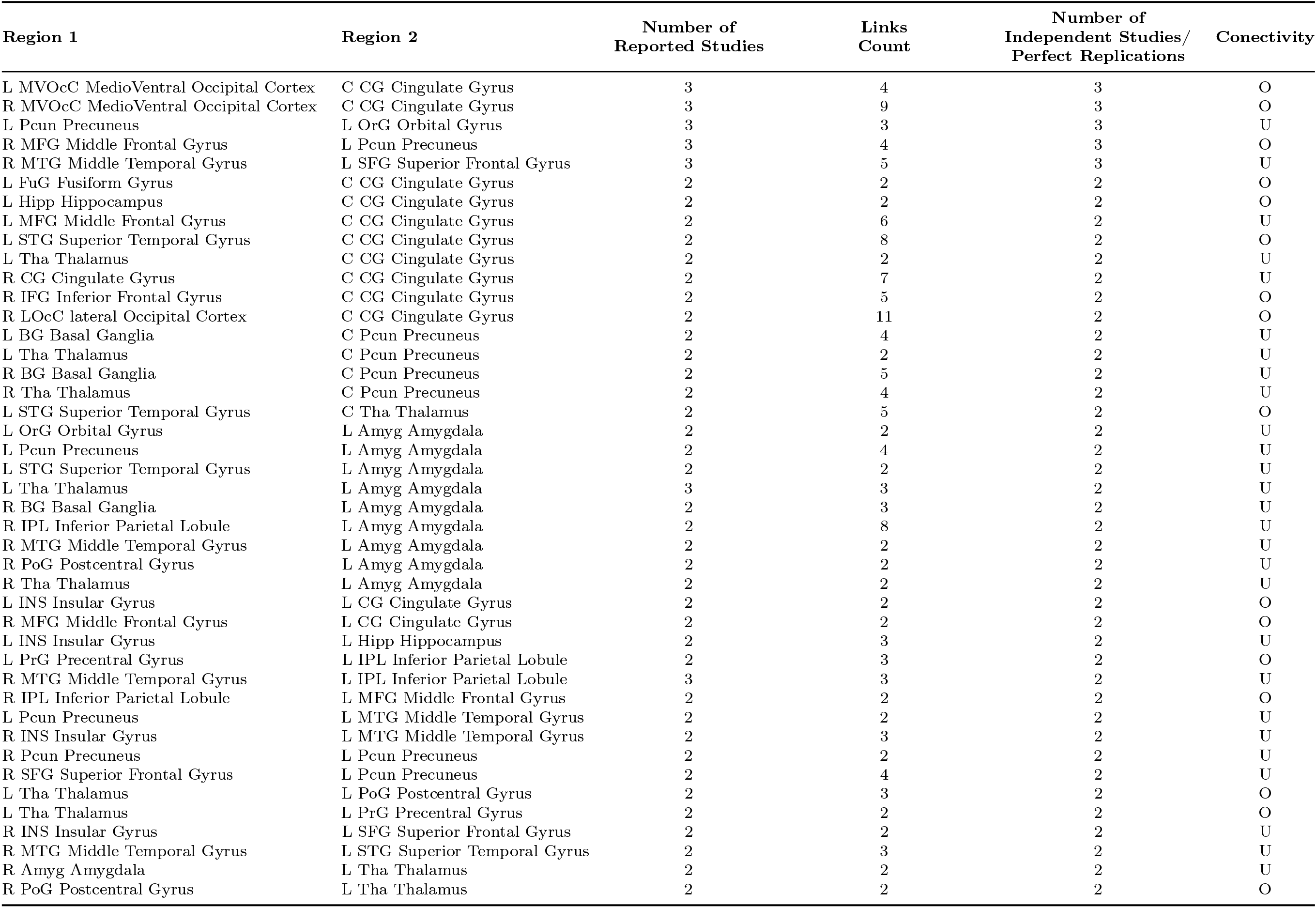
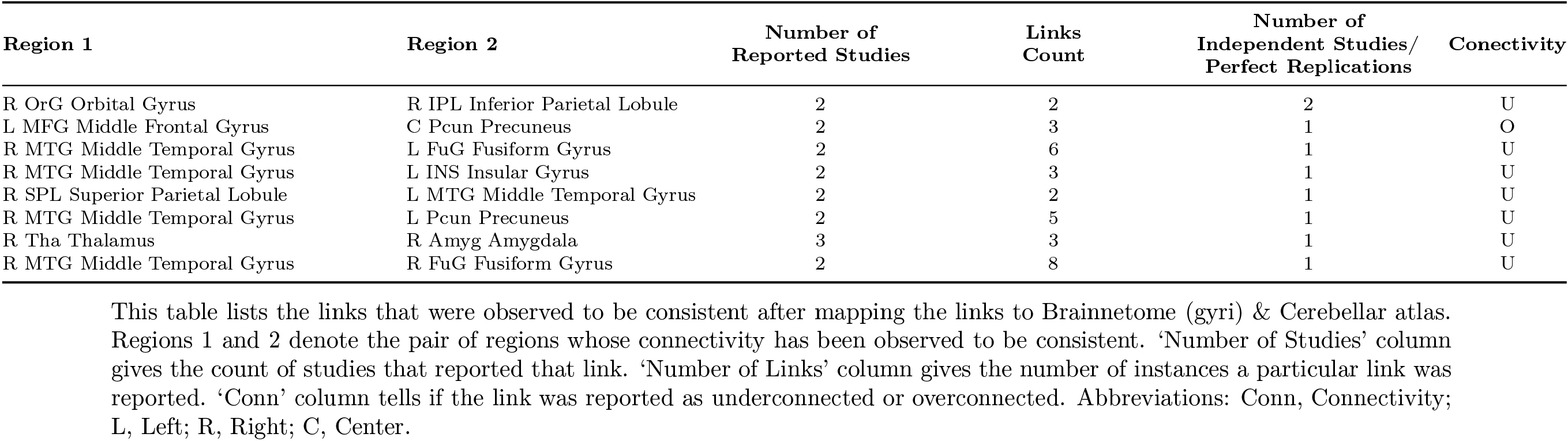
Consistent Autism Altered Functional Connectivity Links (Brainnetome (Gyri) & Cerebellar Atlases)

**Table 8.**
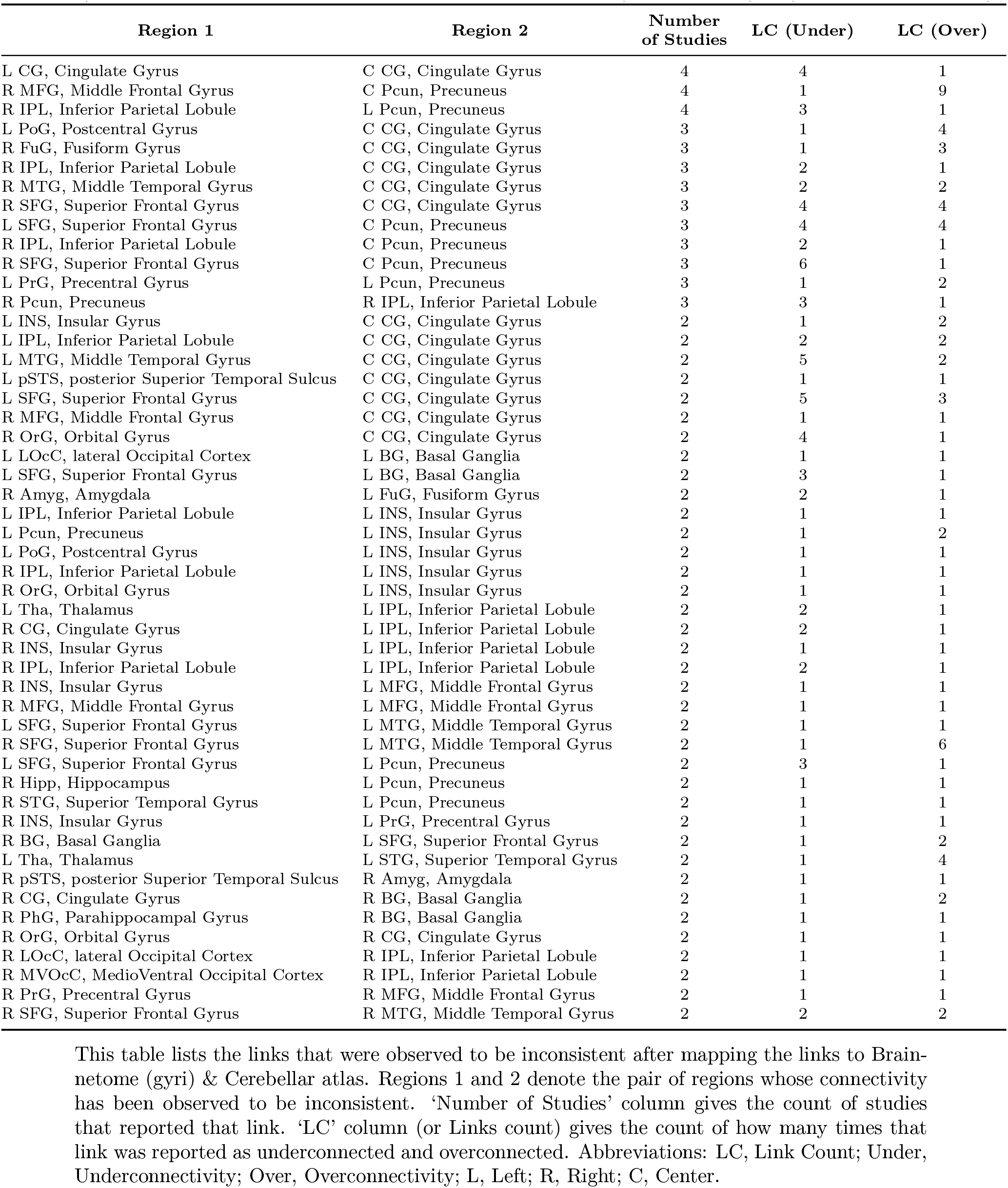
Inconsistent Autism Altered Functional Connectivity Links (Brainnetome (Gyri) & Cerebellar Atlases)

### 3.5. Lobe and the Subcortical Nuclei Level Autism Altered Functional Connectivity Patterns

To examine the overall altered patterns of functional connectivity, the AACLs were mapped to the following five lobes and the subcortical neuclei given by the Brainnetome atlas: frontal lobe, temporal lobe, parietal lobe, limbic lobe, occipital lobe, and subcortical nuclei. The parahippocampal gyrus was included in limbic lobe as mentioned in (Kandel et al., 2000) instead of temporal lobe as in Brainnetome atlas. The cerebellar atlas mask was also used as another lobe making the total count of lobes to seven. Table 9 summarizes the 932 autism altered functional connectivity links extracted from the 44 studies reviewed at lobe level granularity.

**Table 9.** Autism Altered Functional Connectivity Links (Brainnetome (Lobe) & Cerebellar Atlases)

| Lobe 1 | Lobe 2 | Number of Studies | LC (Under) | LC (Over) |
| --- | --- | --- | --- | --- |
| Parietal Lobe | Frontal Lobe | 17 | 48 | 45 |
| Temporal Lobe | Subcortical Nuclei | 16 | 23 | 30 |
| Subcortical Nuclei | Parietal Lobe | 15 | 55 | 28 |
| Subcortical Nuclei | Frontal Lobe | 12 | 20 | 22 |
| Temporal Lobe | Frontal Lobe | 11 | 28 | 28 |
| Subcortical Nuclei | Limbic Lobe | 11 | 19 | 13 |
| Temporal Lobe | Parietal Lobe | 11 | 50 | 3 |
| Parietal Lobe | Parietal Lobe | 10 | 22 | 14 |
| Limbic Lobe | Frontal Lobe | 9 | 31 | 29 |
| Subcortical Nuclei | Subcortical Nuclei | 9 | 20 | 4 |
| Parietal Lobe | Limbic Lobe | 8 | 19 | 21 |
| Temporal Lobe | Limbic Lobe | 8 | 12 | 26 |
| Insular Lobe | Frontal Lobe | 6 | 14 | 9 |
| Parietal Lobe | Insular Lobe | 6 | 6 | 11 |
| Subcortical Nuclei | Insular Lobe | 6 | 11 | 3 |
| Parietal Lobe | Occipital Lobe | 6 | 6 | 3 |
| Subcortical Nuclei | Cerebellum | 5 | 5 | 4 |
| Frontal Lobe | Frontal Lobe | 5 | 19 | 10 |
| Limbic Lobe | Insular Lobe | 5 | 3 | 7 |
| Limbic Lobe | Limbic Lobe | 5 | 17 | 2 |
| Subcortical Nuclei | Occipital Lobe | 5 | 28 | 5 |
| Frontal Lobe | Cerebellum | 4 | 20 | 1 |
| Parietal Lobe | Cerebellum | 4 | 5 | 4 |
| Occipital Lobe | Frontal Lobe | 4 | 4 | 3 |
| Temporal Lobe | Insular Lobe | 4 | 12 | 0 |
| Cerebellum | Cerebellum | 3 | 5 | 6 |
| Temporal Lobe | Cerebellum | 3 | 4 | 5 |
| Occipital Lobe | Limbic Lobe | 3 | 0 | 30 |
| Temporal Lobe | Occipital Lobe | 3 | 6 | 2 |
| Temporal Lobe | Temporal Lobe | 3 | 20 | 0 |
| Limbic Lobe | Cerebellum | 2 | 7 | 6 |
| Insular Lobe | Cerebellum | 1 | 7 | 1 |
| Occipital Lobe | Cerebellum | 1 | 1 | 0 |
| Insular Lobe | Insular Lobe | 1 | 0 | 1 |
| Occipital Lobe | Insular Lobe | 1 | 4 | 0 |

This table lists the links observed after mapping the links to Brainnetome (Lobe) & Cerebellar atlas. Lobe 1 and 2 denote the pair of lobes whose connectivity has been reported. ‘Number of Studies’ column gives the count of studies that reported that link. ‘LC’ column (or Links count) gives the count of how many times that link was reported as underconnected and overconnected. Abbreviations: LC, Link Count; Under, Underconnectivity; Over, Overconnectivity.

This information is graphically represented in Figure 9 describing the unique links that are replicated in at least five studies. The highest number of reported links (93) appeared between parietal and frontal lobes spanning seventeen studies with approximately half (48) reporting underconnectivity and another half (45) reporting overconnectivity in ASD. Next highest (83) are links between subcortical nuclei and parietal lobe appearing in 15 studies with 55 underconnectivity and 28 overconnectivity links. This is followed by 60 links between limbic and frontal lobe spanning nine studies with 31 underconnected links and 29 overconnected links.

**Figure 9:**
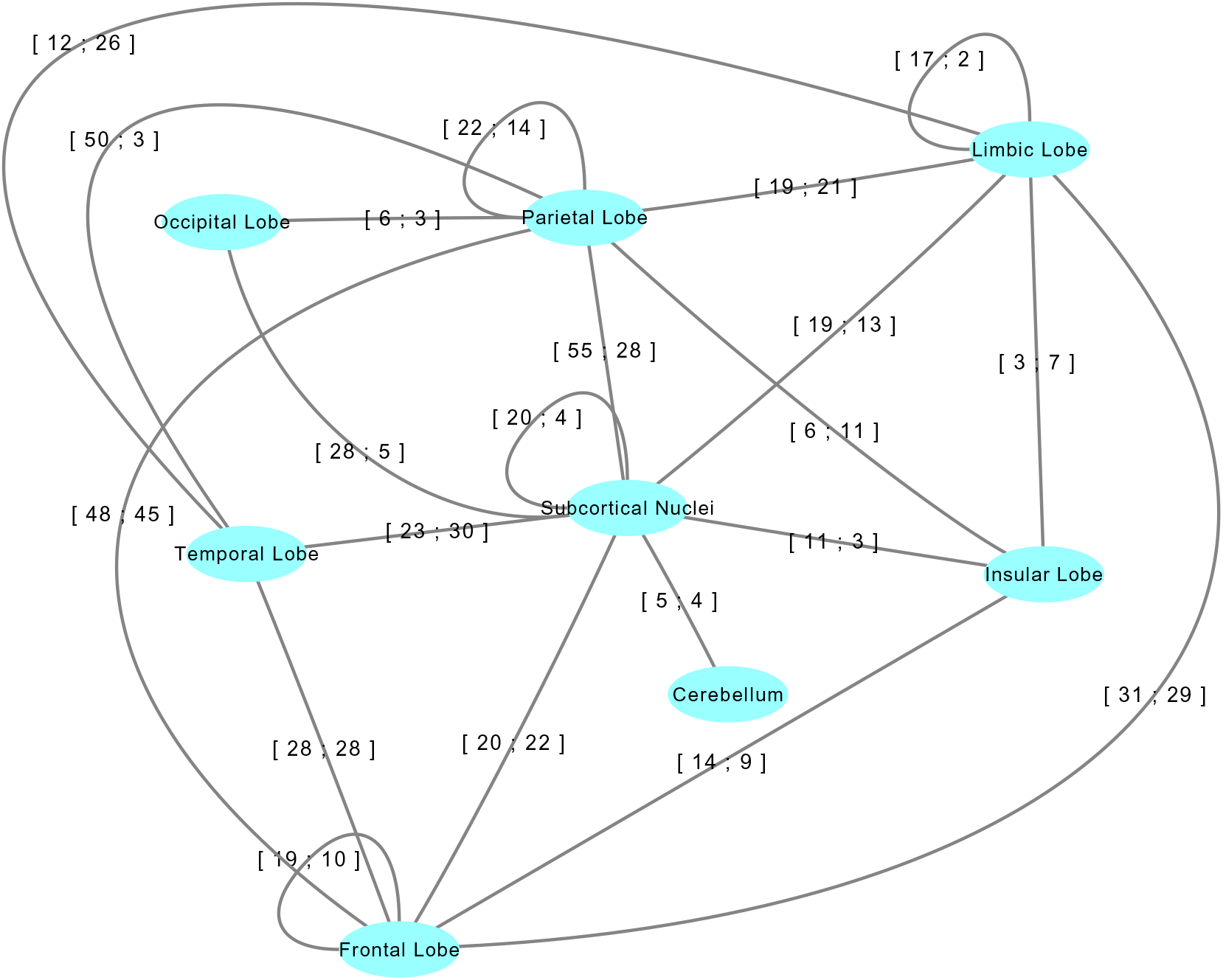
Lobe Level mapping of links that are replicated by at least five studies. The edge label tuple represents the number of under and overconnected AAFLs in the same order as displayed.

Interestingly Table 9 also shows several interesting asymmetries in under and overconnectivity. For example, out of 53 links between temporal and parietal lobes reported by 11 studies, 50 are underconnected. Similarly nine studies have reported 20 underconnectivity and only 4 overconnectivity links among regions of subcortical nuclei, five studies have reported 28 underconnected and 5 overconnected links from subcortical nuclei and occipital lobe, cerebellum was found to be majorly underconnected with the Frontal lobe with 20 out of 21 links spanning 4 studies reported to be underconnected, underconnectivity between insular and temporal lobes with all 12 links of underconnectivity spanning across 4 studies, and 3 studies have found 20 underconnected links among temporal lobes with no overconnected links. Eleven out of 14 links between the insular lobe and subcortical nuclei spanning six studies were found to be underconnected between. This points to the underconenctivity of insula with amygdala, hippocampus, and basal ganglian structures. Overconnectivity was observed across all the 30 links between occipital lobe and limbic lobe spanning across three studies. PCC being a major part of the limbic lobe as well as DMN, points to the overconnectivity between DMN and occipital lobe. Majority of underconnectivity within limbic lobe (17 out of 19 unique links) spanning five studies points to the within DMN under-connectivity.

### 3.6. Network Level Autism Altered Functional Connectivity Patterns

For analyzing the patterns of connectivity of AAFLs at the brain network level, a brain atlas by Schaefer et al. (Schaefer et al., 2017) was employed that parcellates the brain into 100 regions and clusters them into seven networks, namely, Visual, Somatomotor, DMN, Dorsal Attention, Ventral Attention, Limbic, and Control network.

The 932 AAFLs were mapped onto Schaefer brain atlas and Cerebellar atlas. Out of these, 683 links had their endpoints in either the Shaefer brain atlas or cerebellar atlas.

These links have been summarized in Table 10 and graphically represented in Figure 10 with links between functional networks that have been reported in five or more studies.

**Figure 10:**
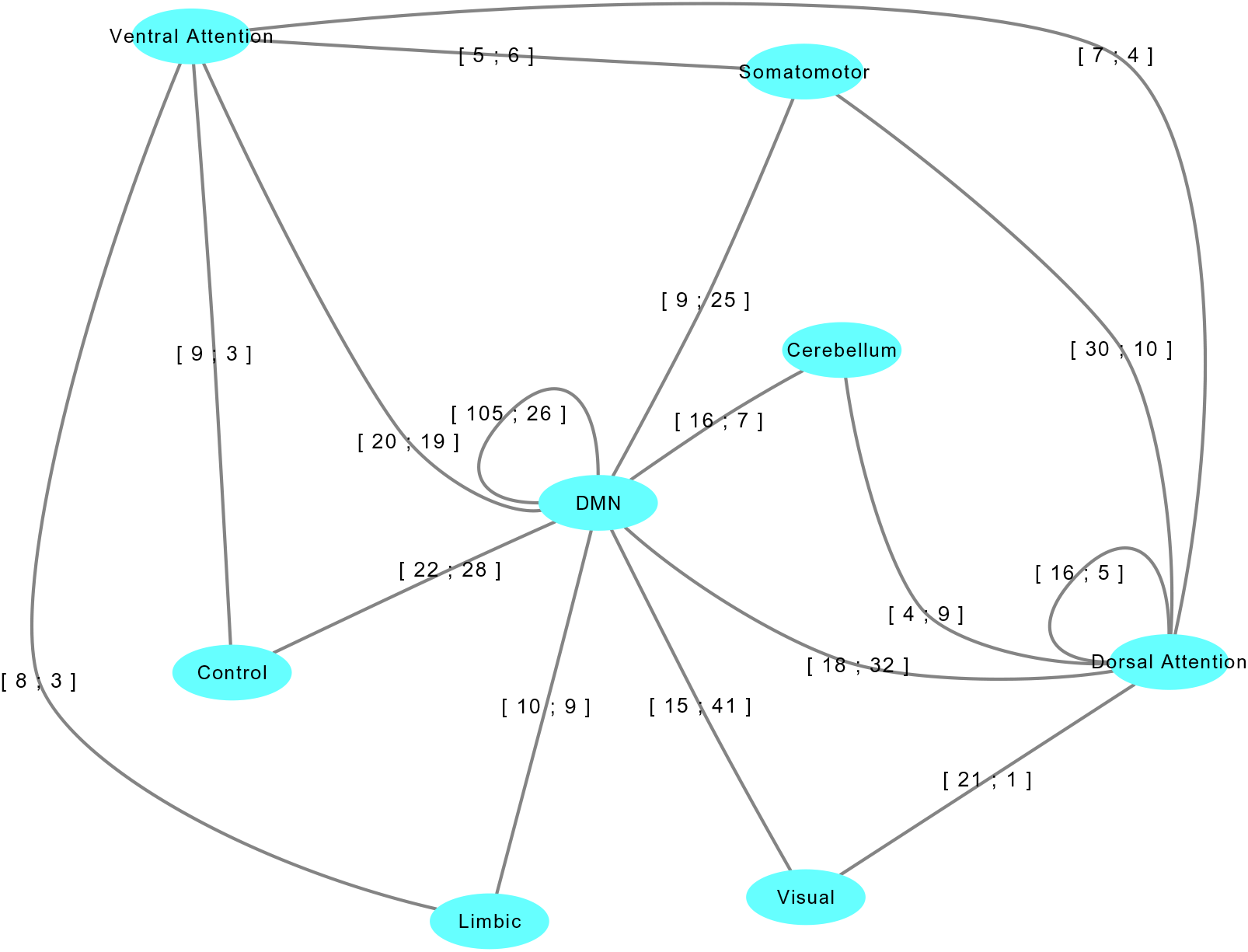
Network Level mapping of links that are replicated by at least five studies. The edge label tuple represents the number of under and overconnected AAFLs in the same order as displayed Abbreviation: DMN, Default Mode Network.

**Table 10.**
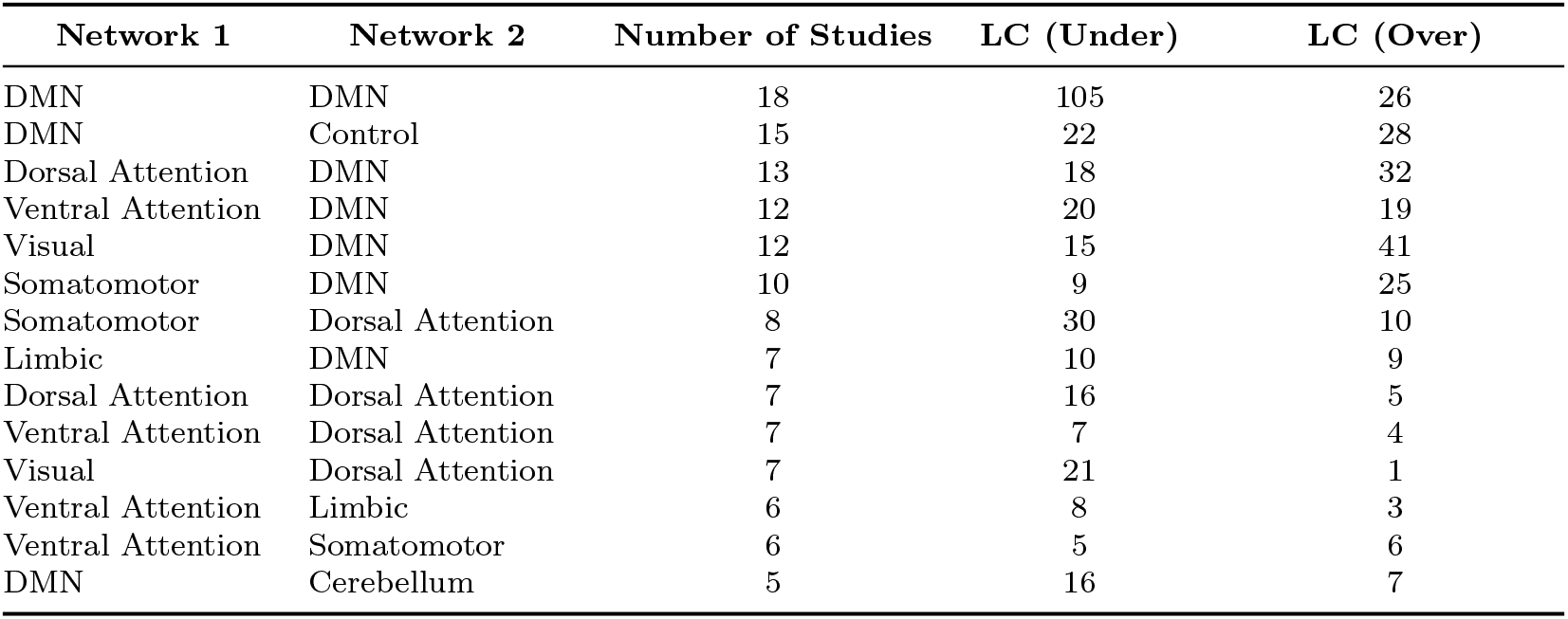
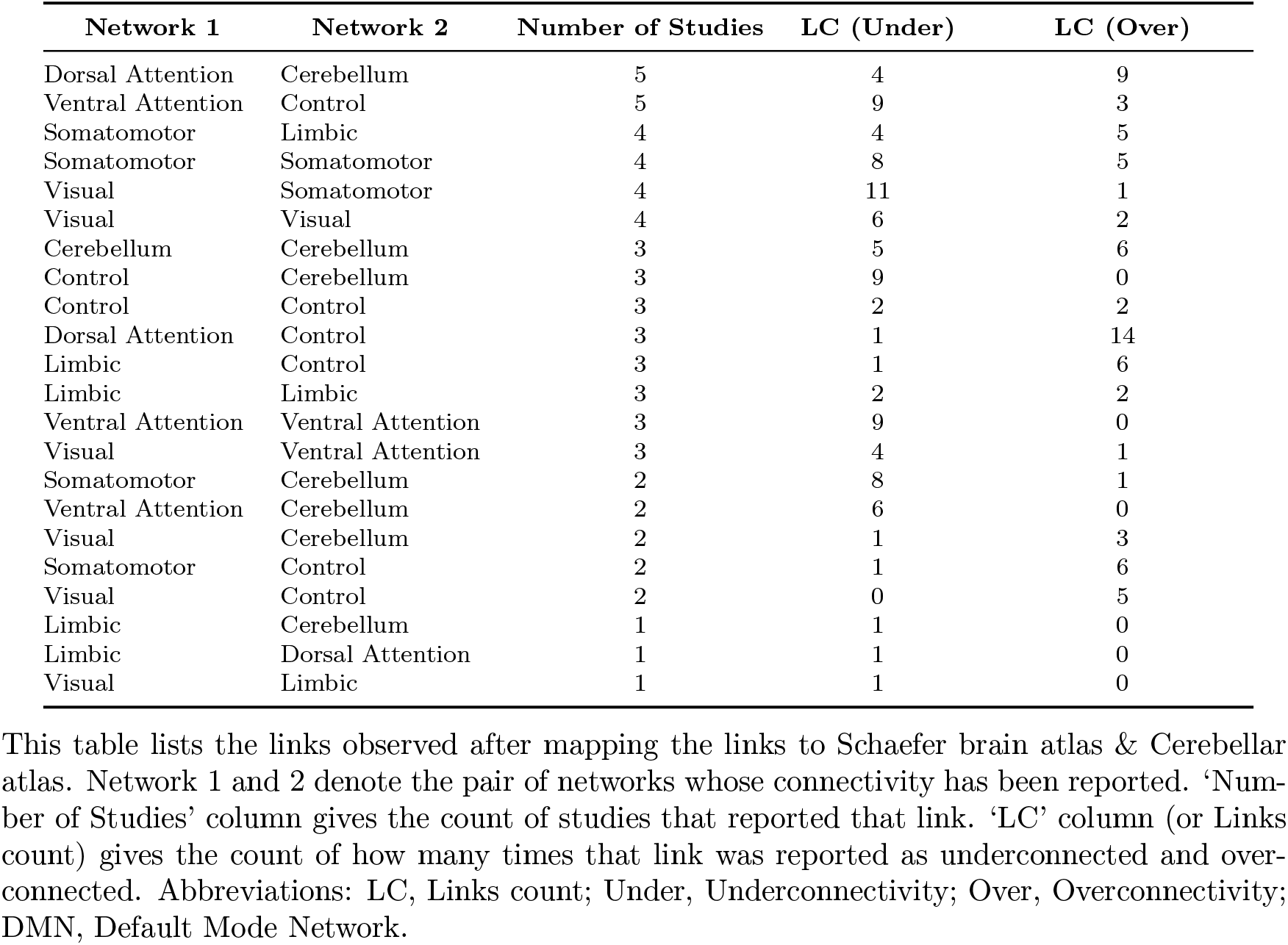
Autism Altered Functional Connectivity Links (Schaefer & Cerebellar Atlases)

| Network 1 | Network 2 | Number of Studies | LC (Under) | LC (Over) |
| --- | --- | --- | --- | --- |
| DMN | DMN | 18 | 105 | 26 |
| DMN | Control | 15 | 22 | 28 |
| Dorsal Attention | DMN | 13 | 18 | 32 |
| Ventral Attention | DMN | 12 | 20 | 19 |
| Visual | DMN | 12 | 15 | 41 |
| Somatomotor | DMN | 10 | 9 | 25 |
| Somatomotor | Dorsal Attention | 8 | 30 | 10 |
| Limbic | DMN | 7 | 10 | 9 |
| Dorsal Attention | Dorsal Attention | 7 | 16 | 5 |
| Ventral Attention | Dorsal Attention | 7 | 7 | 4 |
| Visual | Dorsal Attention | 7 | 21 | 1 |
| Ventral Attention | Limbic | 6 | 8 | 3 |
| Ventral Attention | Somatomotor | 6 | 5 | 6 |
| DMN | Cerebellum | 5 | 16 | 7 |

**Table 10 | continued from previous page**
| Network 1 | Network 2 | Number of Studies | LC (Under) | LC (Over) |
| --- | --- | --- | --- | --- |
| Dorsal Attention | Cerebellum | 5 | 4 | 9 |
| Ventral Attention | Control | 5 | 9 | 3 |
| Somatomotor | Limbic | 4 | 4 | 5 |
| Somatomotor | Somatomotor | 4 | 8 | 5 |
| Visual | Somatomotor | 4 | 11 | 1 |
| Visual | Visual | 4 | 6 | 2 |
| Cerebellum | Cerebellum | 3 | 5 | 6 |
| Control | Cerebellum | 3 | 9 | 0 |
| Control | Control | 3 | 2 | 2 |
| Dorsal Attention | Control | 3 | 1 | 14 |
| Limbic | Control | 3 | 1 | 6 |
| Limbic | Limbic | 3 | 2 | 2 |
| Ventral Attention | Ventral Attention | 3 | 9 | 0 |
| Visual | Ventral Attention | 3 | 4 | 1 |
| Somatomotor | Cerebellum | 2 | 8 | 1 |
| Ventral Attention | Cerebellum | 2 | 6 | 0 |
| Visual | Cerebellum | 2 | 1 | 3 |
| Somatomotor | Control | 2 | 1 | 6 |
| Visual | Control | 2 | 0 | 5 |
| Limbic | Cerebellum | 1 | 1 | 0 |
| Limbic | Dorsal Attention | 1 | 1 | 0 |
| Visual | Limbic | 1 | 1 | 0 |

This table lists the links observed after mapping the links to Schaefer brain atlas & Cerebellar atlas. Network 1 and 2 denote the pair of networks whose connectivity has been reported. ‘Number of Studies’ column gives the count of studies that reported that link. ‘LC’ column (or Links count) gives the count of how many times that link was reported as underconnected and overconnected. Abbreviations: LC, Links count; Under, Underconnectivity; Over, Overconnectivity; DMN, Default Mode Network.

DMN was the most studied brain network. Of the 683 links, 402 links had at least one end-point as a region of DMN. Of the 402 links, there were 131 intra-DMN links (105 underconnected and 26 overconnected), i.e., altered links between DMN regions, and 271 (110 underconnected and 161 overconnected links) extra-DMN links, i.e., altered connectivity links between DMN regions and regions of the brain outside DMN. With 80.15% of total intra-DMN altered connectivity links being underconnected and 59.40% of the total extra-DMN altered connectivity links being overconnected, we conclude that intra-DMN regions are poorly integrated within themselves as well as poorly segregated from the extra-DMN regions. **Considering only the links obtained from original studies, similar trend was observed. 78.66% of DMN-DMN links were under-connected and 61.58% of DMN-others links were over-connected pointing to the existence of weak integration and segregation of DMN.**

The visual network is observed to be majorly overconnected with DMN with 41 unique links of overconnectivity and just 15 unique links of underconnectivity spanning across 12 studies, and majorly underconnected with dorsal attention. Twenty-one out of 22 unique altered functional connectivity links spanning seven studies between visual and dorsal attention network are observed to be underconnectivity links. With thirty out of 40 unique altered functional connectivity links spanning eight studies as underconnectivity links, dorsal attention and somatomotor network are observed to be majorly underconnected.

## 4. Discussion

The AAFC from the 41 publications (44 studies) reviewed with 932 links has been made available as a spreadsheet (see Appendix A.3). This spreadsheet has tabs for (a) study demographics and preprocessing which has information about the study, its demographics, the ASD diagnostic criteria, preprocessing parameters amongst many other related information; (b) the AAFC links tab, that has one one row for each of the altered functional connectivity link along with the representative MNI coordinates for seed region, target region, the names of seed and target regions as reported in the original study, the names of seed and target regions as reported in the AAL Atlas, Harvard-Oxford atlas, the composite Brainnetome (regions) & Cerebellum and, composite Brainnetome (gyri) & Cerebellum atlas, composite Brainnetome (lobe) & Cerebellum atlas, composite Schaefer & Cerebellum atlas, and whether the link was overconnected or underconnected in ASD. All the figures and tables can be generated from this spreadsheet following the methods mentioned in the section 2.7. This extracted data is expected to be a valuable resource for researchers working in this area. The rs-FC research of the past nine years has been summarized quantitatively in the spreadsheet. Future researchers may use this data in combination with other atlases describing regions or brain networks, and the results got using new data to gain more insights into ASD. For example, by aggregating the data over multiple studies, during the network level analysis, the same phenomenon was observed that Joshi and colleagues observed in their study of DMN (Joshi et al., 2017). The intra-DMN regions are poorly integrated

within themselves as well as poorly segregated from the extra-DMN regions. Interestingly, this study was excluded from the review due to not meeting the inclusion criteria of using contiguous regions as seeds, wherein, they took the mean of time courses of multiple regions of DMN.

Study on ADHD individuals by Barber and colleagues (Barber et al., 2015) reported that greater anticorrelation between DMN and occipital region supports better attentional control in typically developing children. Our observation of overconnectivity between DMN and visual network points to the possible explanation of attentional impairment in autism. According to the study by Bebko and colleagues, (Bebko et al., 2015) amygdala-insula rcFC decreases in emotionally dysregulated youth. Underconnectivity observed between the insular lobe and subcortical nuclei especially amygdala might be the possible reason behind emotional impairment of autistic individuals. Temporoparietal junction has been shown to be involved in attention and social interaction (Krall et al., 2015). Underconnectivity observed between temporal and parietal lobes might a possible reason for the impaired attention and social interaction in autism.

The mapping of the reported regions onto the standard brain atlases gave a reproducible way to infer various consistent and inconsistent alterations. Both consistent overconnectivity, as well as underconnectivity in ASD, was observed.

### 4.1. Consistencies

Brainnetome (regions) & Cerebellar atlases mapping of overconnectivity between left precuneus and MFG, and between central precuneus and MFG, and then a reverse inference using the BrainMap database suggests that the differently connected MFG’s region is involved in mental processes of working memory and perception of sensory pain due to injury or emotional disorder, and precuneus is involved in social cognition and explicit memory about events, people, places, etc. This overconnectivity observed between regions exhibiting the mental processes of sensory pain perception, and social cognition might be a reason why autistic children seldom talk with strangers. Mapping of overconnectivity between the thalamus and left STG to Brainnetome atlas shows aberrant functional connectivity of the thalamus with the music and language processing regions of left STG.

Quite different connectivity profiles of two important regions of DMN, namely, precuneus and PCC were observed. Precuneus exhibited a mix of under and overconnectivity with the other brain regions whereas PCC exhibited only overconnectivity with the other brain regions. This shows that representing DMN as the mean of all the regions of DMN or using just one region to represent the complete DMN is not an ideal practice.

### 4.2. Inconsistencies

The majority of inconsistent links was corresponding to the study by Alaerts et al. which analyzed only female subjects (Alaerts et al., 2016) (study ID 40.2). This study reported seven inconsistent links with five studies and just one consistent link between central precuneus and right middle frontal gyrus. Interestingly all these links are reported to be overconnected in this study. This observation points to the possibility of overconnectedness of links in female autistic participants which are generally underconnected in males. Another study that contributed to the major part of the inconsistent links was the study by Eilam et al. (Eilam-Stock et al., 2014). This study reported the maximum number of consistencies as well. One possible reason for a large number of reported consistencies and inconsistencies could be the sheer number of total links reported in this study which is the highest among all the studies. Methodological and participants variability also might have resulted in the inconsistencies. Different preprocessing strategies can influence the group differences. For example, the application of Global Signal Regression (GSR) is debated upon as it alters the group differences (Gotts et al., 2013). Also, head motion affects the functional connectivity (Van Dijk et al., 2012). Regressing out motion parameters is an important step in preprocessing (Friston et al., 1996). Various measures of global motion used by researchers range from the global signal, global correlation, Friston’s 24 motion parameter model (Friston et al., 1996) and the standard six motion parameters model. While most of the studies regressed out the motion parameter model, study by Hogeveen at el. (Hogeveen et al., 2018) regressed out the global correlation of the brain. From the total inconsistent links, this study had contributed two links. These inconsistencies could have been because of the choice of the regressor for removing motion confounds.

A link between left parahippocampal gyrus and right middle temporal gyrus was also observed to be inconsistent. Yerys et al. (Yerys et al., 2015) applied global signal regression (GSR) and observed overconnectivity whereas Alaerts et al. (Alaerts et al., 2016) didn’t apply GSR and observed underconnectivity for the above mentioned link. Another example in which the application of GSR might have caused inconsistency is the link between central ACC and right lingual gyrus which is observed to be inconsistent in two studies - one by Eilam-Stock et al. (Eilam-Stock et al., 2014) that applied GSR and observed overconnectivity and other by Burrows et al. (Burrows et al., 2016) that didn’t apply GSR and reported underconnectivity. It is to be noted that not just the GSR that sets apart the above mentioned two studies. But both the studies are also quite different in terms of preprocessing involved. The study by Abbot et al. (Abbott et al., 2016) regressed the physiological noise using WM and CSF as covariates and used bandpass filter for temporal filtering whereas the study by Weng et al. (Weng et al., 2010) recorded the respiration and heart rate of the participants and used them as covariates. They used just a lowpass filter for temporal smoothing.

Another reason of the inconsistency could be the distance between end points of the inconsistent links. Although some of the inconsistent links may belong to the same pairs of regions, they may be far apart and could represent different functional areas. Table 4 contains the distances of two inconsistent points belonging to the same region. As it may be observed, most of the regions that are inconsistent are far apart. Fifteen of 23 inconsistent links had the total distance (DR_1_ + DR_2_) greater than 20mm till a maximum of 73mm.

### 4.3. Limitations

This survey utilizes a single MNI coordinate to represent an ROI. In practice, different ROIs are of different sizes based on the results and regions investigated. Some studies used huge ROIs such as visual cortex area V1 (Shen et al., 2016). For this review, the whole bilateral ROI was represented with just one coordinate, which may not be an optimal way to represent the region. Moreover, the coordinate when further mapped to Brainnetome brain atlas with finer regions, might give it a different meaning. Another study (Elton et al., 2016) employed bilateral regions of a single seed far apart from the midline, averaged them to get one ROI and did seed-based analysis using that. The present method does not allow this. Therefore, it reports both the left and right ROIs separately assuming that they both have the same altered connectivity.

As discussed before, due to inconsistencies between atlases, the present review relied only on the MNI coordinates reported in the studies. In the case of seed-to-voxels based study, if the seed was defined using an explicitly mentioned atlas and didn’t mention the coordinate, a representative coordinate of the seed was defined. It was defined to be the coordinate of the voxel closest to the center of gravity of that region in the atlas. No study mentioned the name of the atlas that they considered while reporting the results. So, if the representative coordinates of the clusters showing altered connectivity with the seed were not given, they could not be estimated. Therefore these regions were discarded and were not included in the review. Care has been taken in assigning the hemisphere label to the cluster that lies close to the midline. As mentioned in the section 2.7, it is assigned the label named ‘center’. Center label was assigned using the heuristic of the x-coordinate of cluster being less than 6mm, but there could be some regions with large clusters that span both hemispheres and were missed out because of the heuristic.

### 4.4. Reproducibility: Challenges and Way Forward

Finding reproducible results for alterations in rs-FC in ASD was a difficult task. This section describes the difficulties faced and possible methods to make the results more reproducible. Although the difficulties reported are based on our experience of this survey, but we expect this finding to be more generally applicable to fMRI based studies, particular to those investigating rs-FC alterations.

#### Reporting

Some studies just report the region names involved in the altered connectivity, without reporting the atlas used and the coordinates. For example, in the study by Shen and colleagues (Shen et al., 2016), while reporting the results, they did not mention any atlas explicitly. They reported the peak coordinates of the right superior medial frontal gyrus as (12, 44, 19). It corresponded to the right medial frontal gyrus according to Talairach-tournoux atlas and right anterior cingulate cortex according to CA_ML_18_MNIA atlas (AFNI). So, if the region names were used for further analysis, the study may become inconsistent if the region names are reported using a different atlas by another study. This problem is easiest to fix by evolving guidelines standardizing the reporting of the results. These guidelines, using a standardized atlas, may mandate reporting of atlas coordinates for each part of the cluster that spans different brain regions, reporting standardized region name, the corresponding statistical values of the peak voxels (p-value, Z-value or t-value), the values after the correction for multiple comparisons and the methods used for correcting multiple comparisons. It will be ideal to require submission of the final statistical maps in a standardized format (such as DICOM or NIFTI) in the supplementary material along with the publications or at a web platform called Neurovault (Gorgolewski et al., 2015). This will enable a more rigorous meta-studies to be conducted on the results. With the growth of inexpensive storage and computational resources, storing this information on digital platforms should be feasible.

#### Data and code sharing

Sharing of data and code used in generating the results is likely to go a long way in making the research more reproducible. In this light several open source repositories such as ABIDE-I (Di Martino et al., 2013) and II (Di Martino et al., 2017) have done a great service in the advancement of science in this area, and its impact is expected to be felt in coming years. In our review, 14 out of 44 studies considered, used ABIDE-I or 2 data. Other data sharing platforms that host openly accessible data are OpenNeuro (Gorgolewski et al., 2017) and INDI (Biswal et al., 2010). OpenNeuro also incentivizes data sharing by providing data analysis infrastructure and pipelines and hence encourages to make the data and the preprocessing parameters available to the research community so that the studies can be replicated. Source codes of multiple data analysis pipelines such as C-PAC (Sikka et al., 2014) and Porcupine (van Mourik et al., 2018) are openly available on GitHub, and can be used to do reproducible analysis.

#### Diagnostic Criteria

Diagnostic and Statistical Manual of Mental Disorder (DSM) lists down the criteria for classifying someone as having ASD. There have been multiple revisions of DSM, current being the 5*^th^* edition. DSM-V (American Psychiatric Association, 2013) clubs all the diagnostic categories of DSM-IV (American Psychiatric Association, 1994), namely, autism, Aspergers’s syndrome and pervasive developmental disorder not otherwise specified (PDD-NOS) under one umbrella called ASD. Many studies do not mention the exact diagnostic scores behind labeling an individual as having autism, Aspergers’s syndrome or PDD-NOS (according to DSM-IV) or ASD (according to DSM-V). Replicating the results of the studies that investigated the population labeled autistic according to DSM-IV, by using the participant data of studies that investigated population labeled ASD according to DSM-V, is challenging. If the data is shared along with detailed diagnostic scores, then it may be possible to carry out the analysis by pooling the data.

#### Demographics

Different studies investigate participants of different age groups. Child brain evolves and changes structurally (Lenroot and Giedd, 2006) as well as functionally (Anderson, 2002) to an adolescent brain. While doing a systematic review of the kind that is currently presented or a meta-analysis, the variability in the age of the population examined might mask some results that are not consistent across age groups. It was observed that the study by Alaerts et al. which investigated only female participants (Alaerts et al., 2016) (study ID 40.2), was inconsistent with five other studies which had a majority of males. This shows that gender differences may have a major effect on the between-group functional connectivity differences.

#### Preprocessing

Different preprocessing strategies, for example, temporal filtering, global signal regression, have different effects of the functional connectivity alterations. For example, Duan et al. investigated how the functional connectivity alterations change with the change of frequency band while preprocessing (Duan et al., 2017). Gotts et al. showed the effects of regressing out the global brain signal on the functional connectivity alterations. (Gotts et al., 2013).

#### Statistical Analysis Methods

Multiple studies, regress out the confounding variables such as IQ, age and site using linear regression, while many of them do the matching of subjects in the two groups. This may be another reason for finding differences in connectivity alterations. Also, multiple methods are employed by the studies for the corrections of multiple comparisons, namely, False discovery rate (FDR) (Benjamini and Hochberg, 1995), Bonferroni and cluster-extent based thresholding method (Woo et al., 2014), which might also contribute to the difference in findings.

#### Protocol and Scanning

Different protocols employed for scanning such as eyes closed, open or fixated can also lead to group level differences. Other parameters such as TR, number of volumes, scanner sequence (multi-band) can also lead to a difference in the results.

## 5. Conclusions

This systematic review examines all the publications on seed ROI based rs-FC analysis in ASD. It extracts the results of all the prior publications in this area in the form of Altered Functional Connectivity Links (AAFL), represented as two MNI coordinates (of seed region and target cluster/region) and a label representing the alteration in the connectivity in ASD (underconnectivity or overconnectivity) as compared to TD. A meta-analysis could not be carried due to the lack of suitable number of replications. This data is then aggregated to form the Autism Altered Functional Connectome (AAFC) using consistent alterations in functional connectivity in ASD across multiple studies.

Using 41 publications, 932 AAFls were extracted, and 927 got mapped to AAL atlas. These 927 non-unique links translated to 645 unique links, out of which 71 links were replicated (and 38 perfectly replicated). Twenty-two unique inconsistent links and 574 unique links with no replications were found.

These results suggest that the focus of the present research on changes to rs-FC in ASD is on discovery of new results. There are very few attempts to replicate earlier results. With the availability of large ASD related datasets such as ABIDE-I and II, this may change in the future Many studies are beginning to use these datasets (14 out of 44 studies), though, majorly for discovery.

By mapping AAFLs on AAL atlas, we observed that the two important regions of DMN, namely, precuneus and PCC exhibit different altered connectivity profile in ASD. Precuneus is observed to be majorly underconnected and PCC to be overconnected with the other regions of the brain. Network level mapping gave insights into the altered connectivity of DMN. Within-DMN regions are majorly underconnected within themselves and majorly overconnected with extra-DMN regions of the brain. The inconsistent links observed may be explained by the gender distribution of the population investigated as well as the distance between the links.

We identified barriers for replication of the results. With better reporting and publication of statistical maps of the results in a standardized format, many of the difficulties should go away. However, several other factors such as diagnostic criteria, preprocessing or analysis methods will require a much better understanding. More systematic research is needed to develop a better understanding of the subject.

The AAFC has also been made available in the form of a spreadsheet. This may serve as a valuable resource for researchers in this field, as it consolidates all the research results in the last nine years on rs-FC changes (identified using seed ROI based analysis) in ASD at one place in a quantitative, machine as well as human-readable manner.

## Data Availability

Multiple research papers were reviewed in this study and the data/code is compiled whose links are shared.

https://docs.google.com/spreadsheets/d/15X8BZ9_svocLff6G1hNXTBnrfl-dE115emmXhiOVai0/edit?usp=sharing

https://github.com/varun-invent/Autism-survey-connectivity-links-analysis

