## Appendix for "Resting State Functional Connectivity Alterations in Individuals with Autism Spectrum Disorders: A Systematic Review"

### Appendix A. Appendix

*Appendix A.1. Literature Search*

The query used for PubMed database was -

```
(resting[Title/Abstract] OR resting-state[Title/Abstract] OR rest[Title/Abstract]
OR intrinsic[Title/Abstract] OR functional connectivity[Title/Abstract])
AND (fMRI OR functional magnetic resonance imaging OR fcMRI OR functional MRI
OR resting[Title/Abstract])
AND (autism[Title/Abstract] OR autistic[Title/Abstract])
NOT ("task-based"[Title/Abstract] OR "task-based"[Title/Abstract])
AND (connectivity[Title/Abstract])
NOT ("MEG"[Title/Abstract] OR "magnetoencephalography"[Title/Abstract]
OR "independent component analysis"[Title/Abstract] NOT ("seed based"[Title/Abstract]
OR "seed-based"[Title/Abstract]))
```

This resulted in 347 Articles on $9^{th}$April 2018. There were some studies that applied ICA but also did seed-based analysis on anatomically defined regions. So, in order to include these studies double negation was used in the query string.

*Appendix A.1.1. Query Description*

The search query consisted of conjunction of 5 sub-queries.

1. The search terms related to functional connectivity and resting state were - “resting”, “resting-state”, “rest”, “intrinsic” and “functional connectivity”.

   ```
   (resting[Title/Abstract] OR resting-state[Title/Abstract] OR rest[Title/Abstract]
   OR intrinsic[Title/Abstract] OR functional connectivity[Title/Abstract])
   ```

2. The search terms related to fMRI were - “fMRI”, “functional magnetic resonance imaging”, “fcMRI” and “functional MRI”. Again, to make sure the result of this query includes only resting state studies, another search term - “resting”, was included

   ```
   AND (fMRI OR functional magnetic resonance imaging OR fcMRI OR functional MRI
   OR resting[Title/Abstract])
   ```

3. The search terms related to autism were - “autism” and “autistic”.

   ```
   AND (autism[Title/Abstract] OR autistic[Title/Abstract])
   ```

4. The search term “connectivity” was included to get studies that were based on connectivity.

   ```
   AND (connectivity[Title/Abstract])
   ```

5. To exclude the task-based studies the following terms with NOT conjunction - “task-based”, “task-based” were included.

   ```
   NOT (``task based''[Title/Abstract] OR ``task-based''[Title/Abstract])
   ```

6. To exclude the studies related to ICA and MEG the following terms were included with NOT conjunction - “MEG”, “magnetoencephalography”, “independent component analysis”. But to include studies that have used ICA but also have done seed based analysis, another NOT conjunction was added within the current query with terms - “seed based” and “seed-based”.

```
NOT (‘‘MEG’’[Title/Abstract] OR ‘‘magnetoencephalography’’[Title/Abstract]
OR ‘‘independent component analysis’’[Title/Abstract]
NOT (‘‘seed based’’[Title/Abstract] OR ‘‘seed-based’’[Title/Abstract]))
```

*Appendix A.2. Connectomes*

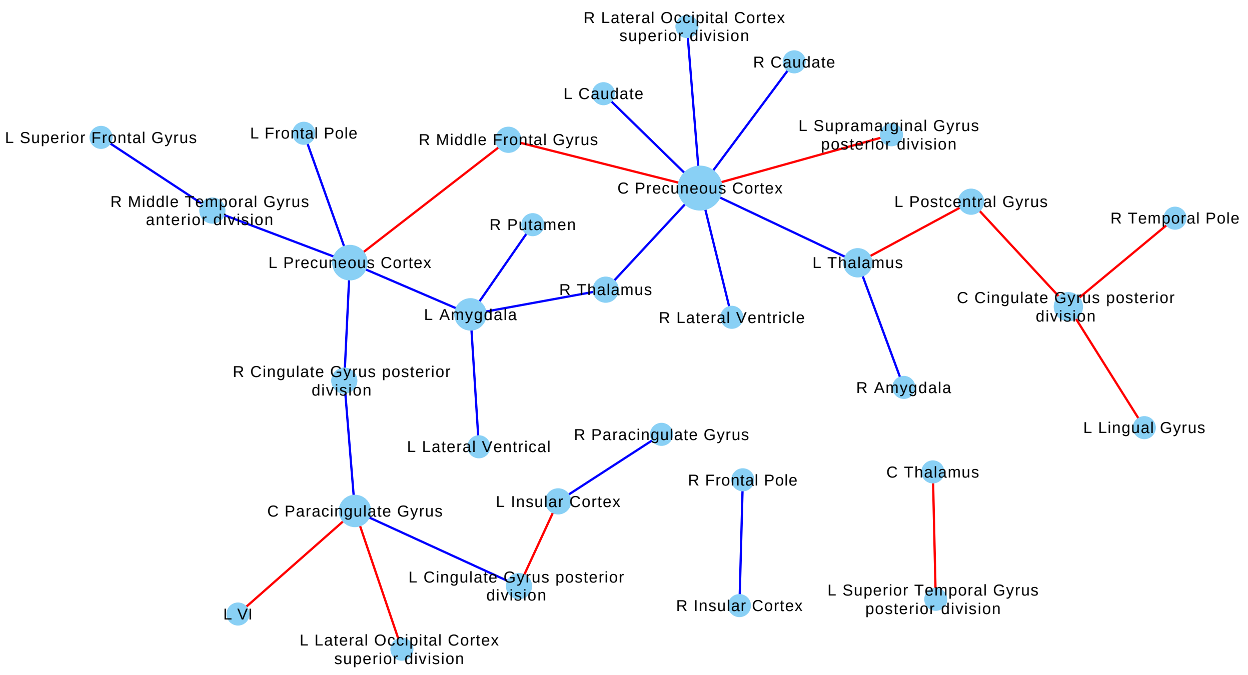


Figure A.11: Connectivity Graph(Harvard Oxford & Probabilistic Cerebellar Atlas): Blue edges represent under-connectivity (i.e. Autistic < TD) and red edges represent over-connectivity. Node size is proportional to the degree of the node. Edge width is proportional to the number of studies with which the link is consistent. Abbreviations: L, Left; R, Right; C, Center.

**Table A.11 | Consistent Autism Altered Functional Connectivity Links (Harvard Oxford & Cerebellar Atlases)**

| Region1 | Region2 | Number of Reported Studies | Links Count | Number of Independent Studies/ Perfect Replications | Conectivity |
|---|---|---|---|---|---|
| L Lingual Gyrus | C Cingulate Gyrus posterior division | 2 | 2 | 2 | O |
| L Postcentral Gyrus | C Cingulate Gyrus posterior division | 2 | 3 | 2 | O |
| R Temporal Pole | C Cingulate Gyrus posterior division | 2 | 2 | 2 | O |
| L Cingulate Gyrus posterior division | C Paracingulate Gyrus | 2 | 2 | 2 | U |
| L Lateral Occipital Cortex superior division | C Paracingulate Gyrus | 2 | 3 | 2 | O |
| L VI | C Paracingulate Gyrus | 2 | 2 | 2 | O |
| R Cingulate Gyrus posterior division | C Paracingulate Gyrus | 2 | 3 | 2 | U |
| L Caudate | C Precuneous Cortex | 2 | 3 | 2 | U |
| L Middle Frontal Gyrus | C Precuneous Cortex | 2 | 3 | 1 | O |
| L Supramarginal Gyrus posterior division | C Precuneous Cortex | 2 | 2 | 2 | O |
| L Thalamus | C Precuneous Cortex | 2 | 2 | 2 | U |
| R Caudate | C Precuneous Cortex | 2 | 2 | 2 | U |
| R Lateral Occipital Cortex superior division | C Precuneous Cortex | 2 | 2 | 2 | U |
| R Lateral Ventricle | C Precuneous Cortex | 2 | 2 | 2 | U |
| R Middle Frontal Gyrus | C Precuneous Cortex | 2 | 6 | 2 | O |
| R Thalamus | C Precuneous Cortex | 2 | 3 | 2 | U |
| L Superior Temporal Gyrus posterior division | C Thalamus | 2 | 2 | 2 | O |
| L Lateral Ventrical | L Amygdala | 2 | 2 | 2 | U |
| L Precuneous Cortex | L Amygdala | 2 | 3 | 2 | U |
| L Thalamus | L Amygdala | 2 | 2 | 1 | U |
| R Putamen | L Amygdala | 2 | 2 | 2 | U |
| R Thalamus | L Amygdala | 2 | 2 | 2 | U |
| R Middle Temporal Gyrus temporooccipital part | L Central Opercular Cortex | 2 | 2 | 1 | U |
| L Insular Cortex | L Cingulate Gyrus posterior division | 2 | 2 | 2 | O |
| L Precuneous Cortex | L Frontal Pole | 2 | 5 | 2 | U |
| R Middle Temporal Gyrus temporooccipital part | L Insular Cortex | 2 | 2 | 1 | U |
| R Paracingulate Gyrus | L Insular Cortex | 2 | 2 | 2 | U |
| L Thalamus | L Postcentral Gyrus | 2 | 3 | 2 | O |
| R Cingulate Gyrus posterior division | L Precuneous Cortex | 2 | 2 | 2 | U |
| R Middle Frontal Gyrus | L Precuneous Cortex | 2 | 2 | 2 | O |
| R Middle Temporal Gyrus anterior division | L Precuneous Cortex | 2 | 5 | 2 | U |
| R Middle Temporal Gyrus anterior division | L Superior Frontal Gyrus | 2 | 3 | 2 | U |
| L Thalamus | L Superior Temporal Gyrus posterior division | 2 | 2 | 1 | O |
| R Middle Temporal Gyrus temporooccipital part | L Temporal Occipital Fusiform Cortex | 2 | 3 | 1 | U |
| R Amygdala | L Thalamus | 2 | 2 | 2 | U |
| R Thalamus | R Amygdala | 3 | 3 | 1 | U |
| R Insular Cortex | R Frontal Pole | 2 | 3 | 2 | U |
| R Temporal Occipital Fusiform Cortex | R Middle Temporal Gyrus temporooccipital part | 2 | 7 | 1 | U |
| R Thalamus | R Precentral Gyrus | 2 | 2 | 1 | O |

**Table A.11 continued from previous page**

| Region1 | Region2 | Number of Reported Studies | Links Count | Number of Independent Studies/ Perfect Replications | Conectivity |
|---|---|---|---|---|---|

This table lists the links that were observed to be consistent after mapping the links to Harvard Oxford (cortical & subcortical) & Cerebellar atlas. Regions 1 and 2 denote the pair of regions whose connectivity has been observed to be consistent. ‘Number of Studies’ column gives the count of studies that reported that link. ‘Number of Links’ column gives the number of instances a particular link was reported. ‘Conn’ column tells if the link was reported as underconnected or overconnected. Abbreviations: Conn, Connectivity.

**Table A.12 | Inconsistent Autism Altered Functional Connectivity Links (Harvard Oxford & Cerebellar Atlases)**

| Region 1 | Region 2 | Number of Studies | LC (Under) | LC (Over) |
|---|---|---|---|---|
| L Postcentral Gyrus | L Middle Temporal Gyrus, temporooccipital part | 2 | 7 | 1 |
| L Superior Frontal Gyrus | C Precuneous Cortex | 2 | 2 | 4 |
| R Frontal Pole | C Precuneous Cortex | 4 | 3 | 3 |
| L Temporal Pole | C Cingulate Gyrus, posterior division | 2 | 1 | 4 |
| R Frontal Pole | C Cingulate Gyrus, posterior division | 2 | 1 | 4 |
| L Superior Frontal Gyrus | C Paracingulate Gyrus | 2 | 3 | 2 |
| R Lateral Occipital Cortex, superior division | C Paracingulate Gyrus | 2 | 1 | 4 |
| R Superior Frontal Gyrus | C Precuneous Cortex | 2 | 3 | 2 |
| L Precentral Gyrus | L Middle Temporal Gyrus, temporooccipital part | 2 | 2 | 3 |
| R Frontal Pole | L Precuneous Cortex | 4 | 2 | 3 |
| L Frontal Pole | C Paracingulate Gyrus | 2 | 2 | 2 |
| L Occipital Pole | L Amygdala | 2 | 3 | 1 |
| R Insular Cortex | L Frontal Pole | 3 | 3 | 1 |
| R Angular Gyrus | L Precuneous Cortex | 4 | 3 | 1 |
| R Precuneous Cortex | R Angular Gyrus | 3 | 3 | 1 |
| L Precuneous Cortex | C Paracingulate Gyrus | 2 | 2 | 1 |
| R Frontal Pole | C Paracingulate Gyrus | 2 | 1 | 2 |
| R Precentral Gyrus | C Paracingulate Gyrus | 2 | 1 | 2 |
| R Temporal Pole | C Paracingulate Gyrus | 2 | 1 | 2 |
| L Precuneous Cortex | L Insular Cortex | 2 | 1 | 2 |
| L Angular Gyrus | C Cingulate Gyrus, posterior division | 2 | 1 | 1 |
| L Lateral Ventrical | C Cingulate Gyrus, posterior division | 2 | 1 | 1 |
| L Parahippocampal Gyrus, anterior division | C Cingulate Gyrus, posterior division | 2 | 1 | 1 |
| L Precuneous Cortex | C Cingulate Gyrus, posterior division | 2 | 1 | 1 |
| R Angular Gyrus | C Paracingulate Gyrus | 2 | 1 | 1 |
| L Precentral Gyrus | L Inferior Frontal Gyrus, pars opercularis | 2 | 1 | 1 |
| L Postcentral Gyrus | L Insular Cortex | 2 | 1 | 1 |
| L Putamen | L Lateral Occipital Cortex, superior division | 2 | 1 | 1 |
| R Angular Gyrus | L Lateral Occipital Cortex, superior division | 2 | 1 | 1 |
| R Insular Cortex | L Lateral Occipital Cortex, superior division | 2 | 1 | 1 |
| L Precuneous Cortex | L Precentral Gyrus | 2 | 1 | 1 |
| R Insular Cortex | L Precentral Gyrus | 2 | 1 | 1 |
| L Superior Frontal Gyrus | L Precuneous Cortex | 2 | 1 | 1 |
| R Parahippocampal Gyrus, posterior division | L Precuneous Cortex | 2 | 1 | 1 |
| L Thalamus | L Supramarginal Gyrus, posterior division | 2 | 1 | 1 |
| R Cingulate Gyrus, anterior division | R Accumbens | 2 | 1 | 1 |
| R Frontal Pole | R Cingulate Gyrus, posterior division | 2 | 1 | 1 |
| R Precentral Gyrus | R Frontal Orbital Cortex | 2 | 1 | 1 |
| R Precentral Gyrus | R Frontal Pole | 2 | 1 | 1 |

Table A.12 | continued from previous page

| Region 1 | Region 2 | Number of Studies | LC (Under) | LC (Over) |
|---|---|---|---|---|

This table lists the links that were observed to be inconsistent after mapping the links to Harvard Oxford (cortical & subcortical) & Cerebellar atlas. Regions 1 and 2 denote the pair of regions whose connectivity has been observed to be inconsistent. 'Number of Studies' column gives the count of studies that reported that link. 'LC' column (or Links count) gives the count of how many times that link was reported as underconnected and overconnected. Abbreviations: Conn, Connectivity; LC, Links count; Under, Underconnectivity; Over, Overconnectivity.

#### *Appendix A.3. Resources*

The spreadsheet given in the URL: `https://docs.google.com/spreadsheets/d/15X8BZ9_svocLff6G1hNXTBnrfl-dE115emmXhiOVai0/edit?usp=sharing` has tabs for (a) study demographics and preprocessing which has information about the study, its demographics, the ASD diagnostic criteria, preprocessing parameters amongst many other related information such as compact representation of the extracted differently connected links; (b) the AAFC links tab, that has one one row for each of the altered functional connectivity link along with representative MNI coordinates for seed region, target region, the names of seed and target regions as reported in the original study, the names of seed and target regions as reported in the AAL Atlas, Harvard-Oxford atlas, the composite Brainnetome (regions) & Cerebellum and, composite Brainnetome (gyri) & Cerebellum atlas, composite Brainnetome (lobe) & Cerebellum atlas, composite Schaefer & Cerebellum atlas, and whether the link was overconnected or underconnected in ASD; and (c) glossary defining all the columns of the spreadsheet. All the figures and tables can be generated from this spreadsheet following the methods mentioned in Section 2.7.

The code used to process the data to derive at various conclusions can be found across two repositories, namely, Autism-survey-connectivity-links-analysis which can be found at `https://github.com/varun-invent/Autism-survey-connectivity-links-analysis`. This repository contains the code that helps in reading the CSV representing the links extracted in the review and finding the consistent and inconsistent links after mapping it to various standard atlases.
